# Microglia-specific polygenic risk stratification reveals distinct pathological subtypes in Alzheimer’s disease

**DOI:** 10.1101/2025.10.06.25336925

**Authors:** Masataka Kikuchi, Akinori Miyashita, Yu Hirota, Norikazu Hara, Mai Hasegawa, Yasufumi Sakakibara, Michiko Sekiya, Kouichi Ozaki, Shumpei Niida, Maho Morishima, Yuko Saito, Shigeo Murayama, Koichi M Iijima, the Japanese Alzheimer’s Disease Neuroimaging Initiative, Takeshi Ikeuchi

**Affiliations:** Department of Molecular Genetics, Brain Research Institute, Niigata University, Niigata, Japan; Department of Neurogenetics, Center for Development of Advanced Medicine for Dementia, National Center for Geriatrics and Gerontology, Aichi, Japan; Department of Experimental Gerontology, Graduate School of Pharmaceutical Sciences, Nagoya City University, Aichi, Japan; Medical Genome Center, Research Institute, National Center for Geriatrics and Gerontology, Aichi, Japan; RIKEN Center for Integrative Medical Sciences, Kanagawa, Japan; Research Institute, National Center for Geriatrics and Gerontology, Aichi, Japan; Department of Neuropathology (the Brain Bank for Aging Research), Tokyo Metropolitan Institute for Geriatrics and Gerontology, Tokyo, Japan; Brain Bank for Neurodevelopmental, Neurological and Psychiatric Disorders, United Graduate School of Child Development, Osaka University, Osaka, Japan

**Keywords:** Alzheimer’s disease, polygenic risk score, microglia, cell-type specificity, spatial transcriptomics, single-nucleus RNA-seq, disease heterogeneity

## Abstract

**INTRODUCTION:** Alzheimer’s disease (AD) is a heterogeneous neurodegenerative disorder in which numerous common variants collectively influence risk. Polygenic risk scores (PRSs) quantify this genetic predisposition, but conventional PRS approaches aggregate biologically diverse variants, potentially masking cell type–specific contributions.

**METHODS:** We developed PRSs for major brain cell types and evaluated their performance in two genomic datasets (n = 4,678 and n = 76). AD cases were stratified by microglia-specific PRS, followed by bulk-brain RNA-seq (n = 16), single-nucleus RNA-seq (n = 15), and spatial transcriptomics of postmortem brains.

**RESULTS:** The microglia-specific PRS most robustly captured AD genetic risk. High–microglia PRS cases exhibited dystrophic microglia with elevated ribosomal gene expression, whereas low–microglia PRS cases exhibited lipid-associated microglia.

**DISCUSSION:** Microglia-focused PRS resolves molecular heterogeneity in AD and provides a framework for the use of cell type–specific genetic models in precision medicine.

## 1. BACKGROUND

Alzheimer’s disease (AD) is driven by a combination of environmental and genetic factors. Unlike disorders caused by single-gene mutations, AD displays a highly polygenic architecture, in which numerous genetic variants with individually small effects collectively contribute to disease susceptibility. These features stand in sharp contrast to early-onset monogenic disorders, which are driven by a single high-impact mutation, whereas late-onset diseases such as AD arise from the cumulative influence of multiple common variants [1].

Polygenic risk scores (PRSs) have emerged as robust tools for quantifying an individual’s genetic susceptibility to disease by aggregating the effects of multiple genetic variants. They offer considerable potential for stratifying patients according to genetic risk and for enabling early interventions before the onset of clinical symptoms [2]. In AD, numerous ancestry-specific PRS models have been developed, with initial efforts focusing on European and North American populations [3, 4]. Subsequent studies have expanded their applicability to diverse ancestral groups [5–7]. Notably, PRSs derived from European-ancestry AD susceptibility loci have maintained strong predictive performance in non-European populations, including Asian cohorts. Beyond risk prediction, these scores are correlated with cerebrospinal fluid biomarker profiles and provide valuable insights into the timing of disease onset [8].

However, late-onset AD is frequently accompanied by other age-related disorders, contributing to clinical and biological heterogeneity that complicates diagnosis. This underlying variability can influence the interpretation of biomarker measurements and the effectiveness of therapeutic interventions, emphasizing the need for more precise patient stratification. Stratifying individuals according to genetic profiles prior to clinical progression could advance the realization of precision medicine and enable tailored preventive or therapeutic strategies. Furthermore, because AD-specific PRSs appear to predict AD dementia more accurately than broader diagnostic categories such as dementia or Alzheimer’s disease and related dementias (ADRD) [8], stratification on the basis of AD-specific PRSs is considered particularly appropriate.

Conventional PRSs are advantageous because they aggregate a large number of single-nucleotide polymorphisms (SNPs) into a single composite risk score. However, the constituent variants often reflect biologically heterogeneous mechanisms, which may reduce their utility for precise patient stratification. For example, among individuals with similarly elevated PRSs, some may harbour genetic vulnerabilities affecting neuronal functions, whereas others may possess innate immune dysfunctions in microglia or astrocytes.

To address this limitation, recent research has increasingly leveraged biological annotations to construct PRS models specific to functional categories or cell types. These approaches include pathway-based PRSs [9–11] and cell type–specific PRSs derived from gene expression signatures characteristic of defined cellular populations [12], both of which have shown the potential to differentiate risk profiles and elucidate underlying disease mechanisms.

Genome-wide association studies (GWASs) of AD have revealed numerous susceptibility loci linked to microglial function [13–15]. Previous single-cell transcriptomic studies have demonstrated that as AD progresses, microglia transition from a homeostatic state to a disease-associated microglial (DAM) state [16, 17]. Building on these findings, recent omics studies focusing on microglia have further refined this classification, revealing distinct DAM subtypes. Notably, lipid droplet–accumulating microglia (LDAMs) represent a subtype detected at elevated levels in the brains of patients with AD [18–21].

These findings suggest that individuals with high microglia-related genetic risk may exhibit altered microglial responses to pathological protein aggregates, including amyloid-β (Aβ) and phosphorylated tau. For example, reduced phagocytic activity has been reported in patients with AD carrying risk variants of TREM2, a microglial membrane receptor that recognizes Aβ [22].

In this study, we constructed cell type–specific PRSs for major brain cell populations, including neurons, microglia, astrocytes, and oligodendrocytes. Among these, the microglia-specific PRS was particularly informative for stratifying AD genetic risk. We then stratified AD patients according to their microglial PRS and applied single-nucleus RNA sequencing (snRNA-seq) and spatial transcriptomics to postmortem brain tissue, identifying molecular pathological differences associated with genetic vulnerability.

Our study introduces a novel framework for genetic risk–based subtyping in AD, enhances our understanding of disease heterogeneity, and highlights potential avenues for precision medicine.

## 2. METHODS

### 2.1 Study Participants

This study included four Japanese cohorts: the postmortem brain (PMB), the clinical (CL), the Japanese Alzheimer’s Disease Neuroimaging Initiative (J-ADNI), and the neuropathology (NP) cohorts. The samples in the PMB and NP cohorts were obtained from the Brain Bank for Aging Research (BBAR) at the Tokyo Metropolitan Institute for Geriatrics and Gerontology. The samples in the CL cohort were obtained from the Japanese from the Japanese Genetic Consortium for Alzheimer Disease (JGSCAD). SNP array genotyping was conducted on 5,178 individuals across the CL, J-ADNI, and NP cohorts, of whom 4,912 passed quality control (QC). Whole-genome sequencing (WGS) was performed on 100 individuals from the PMB cohort, all of whom passed QC. In this study, the CL, J-ADNI, and NP cohorts, which underwent SNP array genotyping, were defined as Cohort 1, while the PMB cohort, which underwent WGS, was defined as Cohort 2. After individuals with mild cognitive impairment (MCI) were removed and duplicates were removed from the WGS dataset, 4,678 participants were included in the SNP-based analyses. AD patients were defined as individuals who were clinically or pathologically diagnosed with AD within each cohort. The demographic characteristics of all the participants are provided in Tables 1 and S1.

### 2.2 Japanese Postmortem Brain Cohort

The PMB cohort consisted of 100 brain donors [23, 24]. Braak stages for neurofibrillary tangles (NFTs) and senile plaques (SPs) were assigned to each donor: Braak NFT stages 0–II (n = 37), III (n = 24), and IV–VI (n = 39); Braak SP stages 0–B (n = 55) and C (n = 45). Pathological controls were defined as donors with SP stage 0 or A and NFT stage 0–II (n = 37), while pathological AD patients were defined as donors with SP stage C and NFT stage IV–VI (n = 39). Donors with Braak NFT stage III, which is indicative of moderate tau pathology (n = 24), were excluded from the case–control analysis. All the PMB donors passed the WGS QC in Section 2.7. A subset of these samples was also used for bulk brain RNA-seq, snRNA-seq, and spatial transcriptomics analyses. Postmortem brain tissues were obtained from the Brain Bank for Aging Research (BBAR) at the Tokyo Metropolitan Institute for Geriatrics and Gerontology. All the brains were diagnosed and scored neuropathologically according to the BBAR protocol.

### 2.3 Japanese Clinical Cohort

The CL cohort comprised 4,093 participants, including 2,124 non-AD controls and 1,969 AD patients [25]. Following the SNP QC procedures described in Section 2.6, 3,843 participants remained for analysis (1,969 non-AD controls and 1,874 AD patients). Probable AD patients were diagnosed according to the criteria of the National Institute of Neurological and Communicative Disorders and Stroke–Alzheimer’s Disease and Related Disorders Association (NINCDS/ADRDA) [26]. Community-dwelling older adults living independently without signs of dementia were recruited as controls.

### 2.4 Japanese Neuropathological Cohort

The NP cohort comprised 577 brain donors [27], including 365 control donors with minimal AD-related pathological changes and 212 case donors with neuropathological features consistent with AD. All AD patients were neuropathologically diagnosed on the basis of the presence of senile plaques and neurofibrillary tangles. No pathological features indicative of other neurodegenerative disorders, such as dementia with Lewy bodies, frontotemporal lobar degeneration, or Parkinson’s disease, were observed. Control donors did not exhibit the characteristic neuropathological hallmarks of AD. As no clinical diagnosis was available for this cohort, the terms “case” and “control” are used herein because they are based solely on neuropathological criteria. After the SNP QC procedures described in Section 2.6 were performed, 565 brain donors remained for analysis (358 controls and 207 cases). Fourteen overlapping samples between the PMB WGS dataset and the NP SNP array dataset were excluded from the SNP-based analyses.

### 2.5 J-ADNI Cohort

The data used in the preparation of this study were obtained from the J-ADNI database deposited in the National Bioscience Database Center Human Database, Japan (Research ID: hum0043.v1, 2016) [28]. This database included cognitively unimpaired participants, participants with late mild cognitive impairment (MCI), and patients with mild AD dementia using criteria consistent with those of the North American ADNI [29]. The J-ADNI was launched in 2007 as a public–private partnership led by principal investigator Takeshi Iwatsubo, MD, with the aim of determining whether serial magnetic resonance imaging, positron emission tomography, other biological markers, and clinical and neuropsychological assessments could be combined to measure the progression of late MCI and mild AD in the Japanese population.

A total of 715 volunteers aged 60–84 years were screened and classified as having late MCI, mild AD, or being cognitively unimpaired. Of these, 537 met the eligibility criteria and were included in the study. Among the enrolled participants, 508 participants underwent SNP array genotyping (non-AD controls, n = 147; MCI, n = 221; AD cases, n = 140). After SNP QC in Section 2.6, 504 participants remained (non-AD controls, n = 145; MCI, n = 220; AD cases, n = 139). For the present study, only non-AD control and AD case data were used.

### 2.6 SNP Array Analysis

Whole blood samples from 4,601 participants in the CL and J-ADNI cohorts, as well as post-mortem frontal cortex samples from 577 donors in the NP cohort, were genotyped using the Infinium Asian Screening Array (Illumina). APOE genotypes were determined for each participant on the basis of haplotypes derived from rs7412 and rs429358, which were genotyped using TaqMan Assays (Applied Biosystems).

We excluded SNPs that (i) had duplicated genomic positions, (ii) had a call rate < 5%, (iii) deviated from Hardy–Weinberg equilibrium in controls (p < 1 × 10⁻⁵), or (iv) had a minor allele frequency < 0.01. For sample-level QC, participants were excluded if they (i) had sex inconsistencies, (ii) showed autosomal heterozygosity deviation (|Fhet| ≥ 0.2), (iii) had < 99% of genotypes called, or (iv) were related (pi-hat > 0.2). Outliers were further removed by principal component analysis (PCA) using reference samples from the 1000 Genomes Project (Phase 3 v5) [30].

Phasing was performed using Eagle v2.4.1 [31], and genotype imputation was conducted with Minimac4 [32] using whole-genome sequencing data from 1,037 participants in the BioBank Japan Project [33] as the reference panel. The above QC procedures were repeated for the imputed SNP markers, and SNPs with poor imputation quality (r² ≤ 0.3) were removed.

The final dataset included SNP genotyping data from 3,843 participants in the CL cohort (1,969 non-AD controls and 1,874 AD patients), 504 participants from the J-ADNI cohort (145 non-AD controls, 220 MCI patients, and 139 AD patients), and 565 brain donors from the NP cohort (358 non-AD controls and 207 AD patients).

### 2.7 Whole-Genome Sequencing Analysis

Genomic DNA was extracted from the postmortem frontal cortex of 100 donors. The DNA was sheared to approximately 350 bp, and sequencing libraries were prepared using the TruSeq DNA PCR-Free Library Prep Kit (Illumina, San Diego, CA, USA). Sequencing was performed on the Illumina HiSeq X Ten system in 151-cycle paired-end mode. Reads were aligned to the human reference genome (GRCh37) using BWA (v0.7.17-r1188) [34], and PCR duplicates were marked and removed using Picard (v2.20.2). Variant calling was performed with GATK (v4.2.0.0) [35] following the GATK4 Best Practices workflow. Individual variants were called using GATK HaplotypeCaller, and joint genotyping across samples was conducted using GATK GenotypeGVCFs. Sample QC was conducted as described for the SNP array analysis, and all the samples passed QC.

### 2.8 Brain Cell Type–Specific Enhancer and Promoter Regions

Brain cell type–specific enhancer and promoter regions were based on the results of Nott *et al.* [36]. These regions were identified from nuclei isolated from neurons, microglia, astrocytes, and oligodendrocytes in the human postmortem cerebral cortex. In brief, cell type–specific nuclear populations were subjected to transposase-accessible chromatin sequencing (ATAC-seq) to identify regions of open chromatin. In addition, H3K27ac and H3K4me3 chromatin immunoprecipitation sequencing (ChIP-seq) were performed to define enhancer and promoter regions, respectively, for each cell type. On the basis of the genomic coordinates provided in the published BED files, we extracted genetic variants located within these regions for downstream analyses. Summary statistics for the enhancer and promoter regions in neurons, microglia, astrocytes, and oligodendrocytes are provided in Table S2.

### 2.9 Calculation of PRS and Prediction Accuracy

The PRS for each individual was calculated as the following weighted sum:

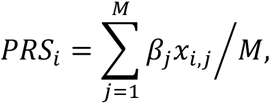

where *PRS_i_* denotes the PRS for individual *i*; *M* is the total number of SNPs included; *β_j_* represents the weight of *SNP_j_*, defined as the effect size from an independent GWAS; and *x_i,j_* is the number of minor alleles of *SNP_j_* in individual *i* (0, 1, or 2). Thus, a higher PRS reflects a greater burden of disease-associated alleles.

SNPs were selected using the clumping and thresholding (C+T) method, which is a widely adopted approach in AD research [37, 38]. PRS calculations were performed with PRSice software [39]. Clumping retained the most strongly associated SNP within each LD block, while correlated SNPs were removed. Thresholding excluded variants with GWAS p values exceeding a given threshold (*p* > *p_T_*). We tested the *p_T_* values (5×10^-8^, 1×10^-6^, 1×10^-5^, 1×10^-4^, 1×10^-3^, 1×10^-2^, 0.05, 0.5, and 1.0) to determine the optimal *p_T_*. SNP effect sizes (*β_j_*) were derived from a large European-ancestry AD GWAS [13].

For cell type–specific PRS construction, only SNPs located within the enhancer or promoter regions of the corresponding cell types (Section 2.8) were considered.

The predictive accuracy was assessed using Nagelkerke’s *R^2^*, which represents the proportion of variance explained in distinguishing non-AD controls from AD patients. This value was computed by comparing a full logistic regression model, including the PRS and the first two components from multidimensional scaling (MDS), to a null model containing only the MDS components: *R^2^* = *R^2^_Full_– R^2^_Null_*. The *p_T_* yielding the highest *R^2^* was selected, and the model constructed at this threshold was designated the best-fit model. All Nagelkerke’s *R^2^* calculations were performed using PRSice software with default parameters [39].

### 2.10 Microglial eQTL Genes

To determine whether the SNPs included in the PRS model influence the expression of specific genes, we utilized the SingleBrain and MiGA3 datasets [40]. The SingleBrain dataset is a meta-analysis of four snRNA-seq datasets from human microglia, whereas MiGA3 is a meta-analysis of three single-cell RNA-seq datasets from human microglia. Genes were defined as microglial eQTL genes if they had a false discovery rate (FDR)–adjusted p value < 0.05 in a random-effects meta-analysis. For the purposes of this study, we used genes identified in either the SingleBrain or MiGA3 datasets.

### 2.11 Enrichment Analysis

Gene functional enrichment analysis was performed using the Metascape platform [41] (https://metascape.org). Gene set enrichment analysis (GSEA) was conducted using the fgsea package on the basis of differential expression comparisons. For the reference data, we used the results obtained from the snRNA-seq dataset generated in this study and applied the FindAllMarkers function (logfc.threshold = 0, test.use = “MAST”) to identify marker genes for each cell type or each microglial subcluster. Genes with avg_log2FC > 0 and p_val_adj < 0.05 were defined as cell type-specific genes or marker genes for the corresponding microglial subcluster. To validate known microglial subclusters, publicly available datasets [20, 21, 42, 43] were used as input data. For Nasu–Hakola disease (NHD) and adult-onset leukoencephalopathy with axonal spheroids and pigmented glia (ALSP), we used signature genes specifically reported in the respective studies [44, 45]. Enrichment of the input gene sets was evaluated using the normalized enrichment score (NES) calculated via the fgsea package.

### 2.12 Bulk brain RNA-seq quantification and differential expression analysis

Total RNA was extracted from frontal-cortex tissue using a TRIzol Plus RNA Purification Kit (Thermo Fisher Scientific, Waltham, MA, USA), and RNA integrity was evaluated on an Agilent 2100 Bioanalyzer (Agilent Technologies, Santa Clara, CA, USA). Bulk RNA-seq libraries were prepared from 500 ng of total RNA using the TruSeq Stranded mRNA Sample Prep Kit (Illumina, San Diego, CA, USA). Libraries were sequenced on an Illumina NextSeq 500 platform, generating paired-end reads (2 × 76 bp). Each library produced an average of ≈ 25 million read pairs. Paired-end RNA-seq reads (FASTQ format) were processed using salmon (version 1.7.0) [46] for transcript quantification. Raw FASTQ files were quantified against a prebuilt human transcriptome index generated from the Ensembl human transcriptome (Ensembl release 104, GRCh38) in quasi-mapping mode with automatic library type detection (-l A) and the –validateMappings option enabled.

Transcript-level quantifications from all samples were imported into R using the tximport package (version 1.10.1) [47]. Transcript-to-gene mapping was performed on the basis of the Ensembl transcript ID–gene ID correspondence table, and gene-level counts were obtained using the summarizeToGene function. For samples with multiple technical replicates, mean counts across replicates were calculated to generate a single expression profile per sample.

Differential expression analysis was conducted with the DESeq2 package (version 1.34.0) [48]. Genes were retained only if they met both criteria: (i) total read count ≥ 10 across all samples and (ii) >0 counts in at least half of the samples in the comparison group. A design formula of ∼ groups was applied in DESeq2. Genes with p values < 0.05 were considered to be differentially expressed.

### 2.13 Isolation of nuclei from frozen brain tissue

Nuclei were isolated from approximately 20 mg of fresh-frozen postmortem brain tissue from 15 individuals (Table S3) using the Minute Single Nucleus Isolation Kit for Neuronal Tissues/Cells (Invent Biotechnologies) according to the manufacturer’s protocol. Isolated nuclei were stained with ReadyCount Blue Nuclear Stain (Thermo Fisher Scientific) and counted on a Countess II FL automated cell counter (Thermo Fisher Scientific).

### 2.14 snRNA-seq processing

Isolated nuclei were processed for snRNA-seq using the Chromium Next GEM Single-Cell 3’ Kit v3.1 (10x Genomics) following the manufacturer’s protocol. Briefly, for each library, approximately 10,000 nuclei were partitioned into a gel bead-in-emulsion on a Chromium Controller instrument (10x Genomics). Reverse transcription and cDNA amplification were performed, and Illumina-compatible adapter sequences were added to the final libraries via PCR. The libraries were subsequently sequenced on an Illumina NextSeq2000 instrument. Raw sequencing data were processed using the Cell Ranger pipeline (v7.0.0; 10x Genomics) [49] for demultiplexing, barcode assignment, read alignment to the human reference genome (GRCh38-2020-A), and gene counting.

Data processing and downstream analyses were conducted using the Seurat package (v5.0.3) [50]. Genes were retained if ≥3 nuclei were detected. Nuclei were excluded if they had <1,000 or >5,000 detected genes or if the mitochondrial RNA content exceeded 5%. Doublets were identified using the cxds_bcds_hybrid function from the scds package [51], and ambient RNA contamination was removed using DecontX [52].

Gene expression UMI counts were normalized using SCTransform [53] with regression of mitochondrial counts, followed by dimensionality reduction with the RunPCA and RunUMAP functions in Seurat. Data integration across samples was performed using IntegrateLayers (method = “HarmonyIntegration”, normalization.method = “SCT”). Cell clustering was carried out using the FindNeighbors and FindClusters functions. Microglial clusters were identified on the basis of *CSF1R* and *CD74* expression, and these clusters were subsequently reclustered. Subclusters expressing nonmicroglial marker genes were removed. Dimensionality reduction and reclustering were repeated on the filtered object with identical parameters, yielding eight microglial subclusters at a clustering resolution of 0.3.

### 2.15 Differential gene expression analysis of the snRNA-seq data

Differential gene expression analysis between groups was conducted using the MAST method implemented in the FindMarkers function of the Seurat package. The analysis was adjusted for sex, age at death, *APOE* ε4 allele dose, and postmortem interval. Genes were considered differentially expressed if they satisfied the thresholds of p_val_adj < 0.05 and |avg_log2FC| > 0.5. Pseudobulk analysis was performed to compare sample-level expression between groups. For each sample, the average expression of each gene within a given cell cluster was computed using the AverageExpression function in Seurat, which is based on the SCT-normalized expression matrix.

### 2.16 Spatial Transcriptomics Analysis Using the NanoString GeoMx Digital Spatial Profiler

The tissue samples were embedded in optimal cutting temperature compound and cryosectioned at a thickness of 5 μm. Sections were fixed in 10% neutral buffered formalin at room temperature for 16 h, washed with PBS, baked at 60 °C for 30 min, dehydrated sequentially in 50%, 70%, and 100% ethanol, and air-dried at room temperature for up to 1 h.

Epitope retrieval and staining were performed using a Leica Bond Autostainer. Heat-induced epitope retrieval was conducted with Epitope Retrieval Solution 2 (Leica, AR9640) at 85 °C for 15 min, followed by treatment with proteinase K (0.1 μg/mL; Thermo Fisher, AM2548) at 37 °C for 10 min. The slides were then hybridized overnight at 37 °C with whole-transcriptome RNA detection probes (NanoString Technologies), each conjugated to a unique photocleavable DNA oligonucleotide tag.

Following hybridization, stringent washes were performed twice for 25 minutes at 37 °C using a solution of equal parts 4× saline-sodium citrate (SSC) buffer and 100% formamide, followed by a final wash in 2× SSC buffer. Nonspecific binding was blocked with Buffer W (NanoString proprietary blocking buffer), and the sections were incubated with the following fluorescently labelled antibodies: anti-GFAP mouse monoclonal antibody (Alexa Fluor 532, 4 μg/mL; Novus Biologicals, NBP1-05197AF532), anti–β-amyloid rabbit monoclonal antibody (Alexa Fluor 594, 3 μg/mL; Cell Signaling Technology, 35363S), and anti-Iba1/AIF-1 rabbit monoclonal antibody (Alexa Fluor 647, 4 μg/mL; Cell Signaling Technology, 78060S). Nuclear counterstaining was performed using SYTO 13 (NanoString, GMX-RNA-MORPHHST-12) in Buffer W for 1 h at room temperature, followed by washing in 2× SSC buffer.

All the samples were profiled using the NanoString GeoMx Digital Spatial Profiler (DSP) platform with the Whole Transcriptome Atlas (WTA) probe panel, and transcript quantification was performed using next-generation sequencing. Two tissue sections were mounted per slide. Eurofins Genomics provided technical services for GeoMx DSP–WTA analysis.

### 2.17 Selection of Regions of Interest (ROIs) and Areas of Interest (AOIs)

In the GeoMx platform, regions of interest (ROIs) were selected on tissue sections based on specific antibody markers. To focus on areas with prominent Aβ deposition, Aβ-positive regions were identified by immunohistochemical staining, and rectangular ROIs of the maximum size (660 μm × 785 μm) were selected from the sections. Within each ROI, target cell populations were further defined as areas of interest (AOIs) according to antibody markers. We focused on microglia and defined AOIs using immunostaining for Aβ, IBA1 (microglia), and GFAP (astrocytes) to minimize astrocyte-derived signal interference as follows:

1. On-plaque microglia: IBA1⁺Aβ⁺GFAP⁻ cells located within Aβ deposition sites.
2. Periplaque microglia: IBA1⁺Aβ⁻GFAP⁻ cells located adjacent to, but not overlapping with, Aβ deposits.

In the non-AD control group, where no Aβ deposition was observed, AOIs were defined as IBA1-positive microglia from randomly selected cortical regions. Although multiple AOIs can be selected from a single ROI in the GeoMx platform, the extensive and dense Aβ deposition in AD brains made it difficult to spatially separate “on-plaque” and “periplaque” microglia within the same ROI. Therefore, each AOI was extracted from a distinct ROI, resulting in a one-to-one correspondence between the ROIs and AOIs.

Following NanoString’s recommendations, a preliminary evaluation was performed using adjacent serial sections to confirm that each ROI/AOI contained the minimum number of nuclei required (≥100) for RNA measurement. On the basis of this assessment, 4–10 ROIs/AOIs per case were selected for each sample to ensure compliance with the minimum cell count requirement (Table S4).

### 2.18 Quality Control and Downstream Analysis of GeoMx DSP–WTA Data

FASTQ files generated from the GeoMx platform were processed using the GeomxTools package to convert sequencing data into DCC files containing raw probe count matrices. AOI-level QC was performed according to NanoString’s recommendations, and AOIs were excluded if they met any of the following criteria: >1000 raw reads, <80% aligned, trimmed, stitched, <50% saturation, >1000 NTC, or <100 nuclei. After QC, the final dataset included six samples, three from the high-PRS AD group and three from the low-PRS AD group (Table S4). Both on-plaque and periplaque AOIs were successfully obtained from two brain samples in each PRS group, with a minimum of two AOIs per sample.

Probes were removed if (i) the geometric mean of the probe counts across all segments divided by the geometric mean of all probe counts for the target was <0.1 or if (ii) they were flagged as outliers in ≥20% of the segments by Grubb’s test. Raw gene counts were calculated as the geometric mean of associated probes. Genes and AOIs with abnormally low signals were excluded on the basis of the limit of quantification (LOQ) derived from the distribution of negative control probes, with both the LOQ standard deviation threshold and minimum value set to 2 (NanoString recommendation). Data were normalized using the Q3 method in GeomxTools.

Differential expression analysis was performed using a linear mixed-effects model with a random intercept for slide identity to account for interslide variability. Genes with an FDR–adjusted p value <0.05 and an absolute effect size (Estimate) >0.5 were considered to be differentially expressed.

### 2.19 RNAscope In Situ Hybridization and Immunohistochemistry

Frozen human postmortem temporal cortex sections (5 μm thick) were mounted on Superfrost Plus slides (Fisherbrand, Cat# 12-550-15) and processed according to the manufacturer’s instructions outlined in the RNAscope^®^ 2.5 LS or BaseScope™ RED Assay combined with the Immunohistochemistry: Integrated Co-Detection Workflow (ICW) on the Leica Bond platform (MK 51-151 TN LS RED with ICW). Enzymatic pretreatment was performed with RNAscope LS Protease IV (ACD Biotechne, Cat# 322140) at room temperature for 15 minutes.

The RNAscope probes included Hs-P2RY12-C2 (ACD Biotechne, Cat# 450398-C2) visualized with Opal 520, Hs-RGS1-C1 (ACD Biotechne, Cat# 438598) visualized with Opal 570, Hs-CXCR4-C3 (ACD Biotechne, Cat# 310518-C3) visualized with Opal 690, and Hs-DUSP1-C4 (ACD Biotechne, Cat# 898258-C4) visualized with Opal 780. In situ hybridization and multiplex fluorescent staining were performed using the RNAscope™ LS Multiplex Fluorescent Reagent Kit v2 (ACD Biotechne, Cat# UM 322800) in combination with the Leica Bond Autostainer, following the LS-4-plex-Ancillary-TechNote-06302017 protocol.

Following hybridization, the slides were incubated with a primary antibody against β-amyloid: β-amyloid (D54D2) XP rabbit monoclonal antibody conjugated to Alexa Fluor 594 (Cell Signaling Technology, Cat# 35363S; 1:50 dilution).

Whole-slide fluorescence imaging was performed using the Phenoptics Vectra Polaris Slide Scanner (Akoya Biosciences) at 20× magnification. Spectral unmixing was conducted using inForm image analysis software to separate target signals from each other and from tissue autofluorescence. Processed images were saved as TIFF files and further analysed using the HALO image analysis platform (Indica Labs).

White matter and vascular-rich regions were manually excluded using the pen tool. The analysed area per sample ranged from 39.0 to 61.7 mm². FISH signals were detected using the FISH-IF v2.2.19 module, which enabled nuclear detection and per-cell copy number quantification. Cytoplasmic regions were estimated for each nucleus with consistent parameters across slides (cyto_radius = 3.96, cell_min_area = 10, cell_max_area = 600).

An Aβ-deposit classifier was developed using HALO AI with the DenseNet V2 algorithm, trained on dozens of manually annotated Aβ deposits per section. The classifier automatically identified the location and area of each Aβ deposit within each tissue section. The shortest distances between Aβ deposits and individual cells were calculated using the spatial analysis module. Eurofins Genomics provided technical services for RNAscope and imaging analysis.

### 2.20 Immunostaining

Formalin-fixed brain tissues were embedded in paraffin wax and cut into 6-μm-thick sections [54]. After deparaffinization, the sections were submerged in Tris-EDTA buffer (pH 9.0), heated at 95–97°C for 20 min, and then cooled to room temperature in the same buffer. The sections were immersed in 70% formic acid for 5 min at room temperature after having been subjected to heat-mediated antigen retrieval and were blocked with PBS containing 5% normal donkey serum, 0.5% bovine serum albumin, and 0.3% Triton X-100, and then incubated with anti-RGS1 (GeneTex, GTX112803, 1/100), anti-Iba1 (Abcam, ab5076, 1/100), anti-CD68 (Abcam, ab955, 1/100) or anti-Aβ (BioLegend, 856502, 1/300) antibodies diluted in PBS containing 3% donkey normal serum, 0.5% bovine serum albumin, and 0.3% Triton X-100 at 4°C for 74 h. After washing the sections with PBS three times, they were incubated with appropriate Alexa Fluor-conjugated secondary antibodies in dilution buffer for 3 h at room temperature in the dark. Finally, they were submerged in 0.1% Sudan Black solution in 70% EtOH for 20 min to quench the autofluorescence. They were mounted with Aqua/Poly-Mount and observed under confocal laser-scanning microscopes.

### 2.21 Image acquisition and analysis

Images were acquired using STELLARIS5 (Leica Microsystems) confocal laser-scanning microscope with 63× objectives. 63× images were deconvolved using Leica Lightning Deconvolution (63× objective; z-axis interval 0.29 µm). All image processing and analysis were performed using the Fiji software as described previously [55]. Z-stack confocal images were reconstructed using maximum intensity projections.

### 2.22 3D image analysis by Imaris

To visualize the colocalization between RGS1 and Iba1 signals, we captured images using a STELLARIS5 confocal microscope with a 63× objective lens and reconstructed these images in 3D using Imaris software (Oxford Instruments). Individual structures were created for the surface element. Iba1 surfaces were selected with a smoothness of 0.0721 μm. For colocalization analyses of RGS1-positive signals with Iba1-positive microglia, cell bodies and processes were reconstructed in 3D, and overlap volumes were calculated.

## 3. RESULTS

### 3.1 Construction and Validation of Cell-Type-Specific PRS Models

We constructed cell type–specific PRS models using SNP genotyping data from Cohort 1 (CL, J-ADNI, and NP cohorts), which included 4,678 Japanese individuals (2,469 non-AD controls and 2,209 AD patients; Table 1, Table S1). Compared with controls, AD patients were on average 0.9 years younger (p = 1.91 × 10⁻⁶, Welch’s t test) and comprised a higher proportion of females and *APOE* ε4 carriers and a lower proportion of *APOE* ε2 carriers (all p < 0.001, χ² test).

**Table 1.** Summary of the participants.

|  | Cohort1 (CL, J-ADNI, and NP cohorts) |  |  | Cohort2 (PMB cohort) |  |  |  |
| --- | --- | --- | --- | --- | --- | --- | --- |
|  | Ctrl | Case | p value | Ctrl | Braak3 | Case | p value |
| N | 2,469 | 2,209 | - | 37 | 24 | 39 | - |
| Age in years, mean ± SE | 75.9 ± 0.138 | 75.0 ± 0.132 | **$1.91 \times 10^{-6a}$** | 75.4 ± 1.225 | 83.2 ± 1.011 | 83.3 ± 1.141 | **$1.31 \times 10^{-6b}$** |
| Sex (M:F) | 1,120:1,349 | 659:1,550 | **$< 2.2 \times 10^{-16c}$** | 24:13 | 13:11 | 16:23 | **0.021<sup>d</sup>** |
| APOEε4 alleles (0:1:2) | 2,056:400:13 | 1,036:965:208 | **$< 2.2 \times 10^{-16c}$** | 25:12:0 | 22:2:0 | 16:13:10 | **$2.01 \times 10^{-5d}$** |
| APOEε2 alleles (0:1:2) | 2,248:215:6 | 2,116:92:1 | **$1.12 \times 10^{-10d}$** | 37:0:0 | 24:0:0 | 39:0:0 | 1.000<sup>d</sup> |

<sup>a</sup>two-sided Welch's t test, <sup>b</sup>one-way ANOVA, <sup>c</sup>Chi-squared test, <sup>d</sup>Fisher's exact test. P values in bold are statistically significant.
Abbreviations: APOE, apolipoprotein E; SE, standard error; ANOVA, analysis of variance.

Using summary statistics from a European-ancestry AD GWAS, we developed eight cell type–specific PRS models incorporating SNPs located in enhancer or promoter regions active in neurons, microglia, astrocytes, or oligodendrocytes. In addition, we developed a conventional PRS model using genome-wide SNPs, referred to as the “standard PRS.”

In Cohort 1, the standard PRS achieved the greatest discriminative power for AD (Nagelkerke’s R² = 0.016, p = 2.25 × 10⁻¹⁴, 45 SNPs), followed by the microglial promoter–based PRS (hereafter referred to as MicroProm PRS) (Nagelkerke’s R² = 0.014, p = 1.11 × 10⁻¹², 24 SNPs) (Figures 1A (left panel) and S1, Table S5). All the models contained fewer than 78 SNPs (Table S5).

**Figure 1.**
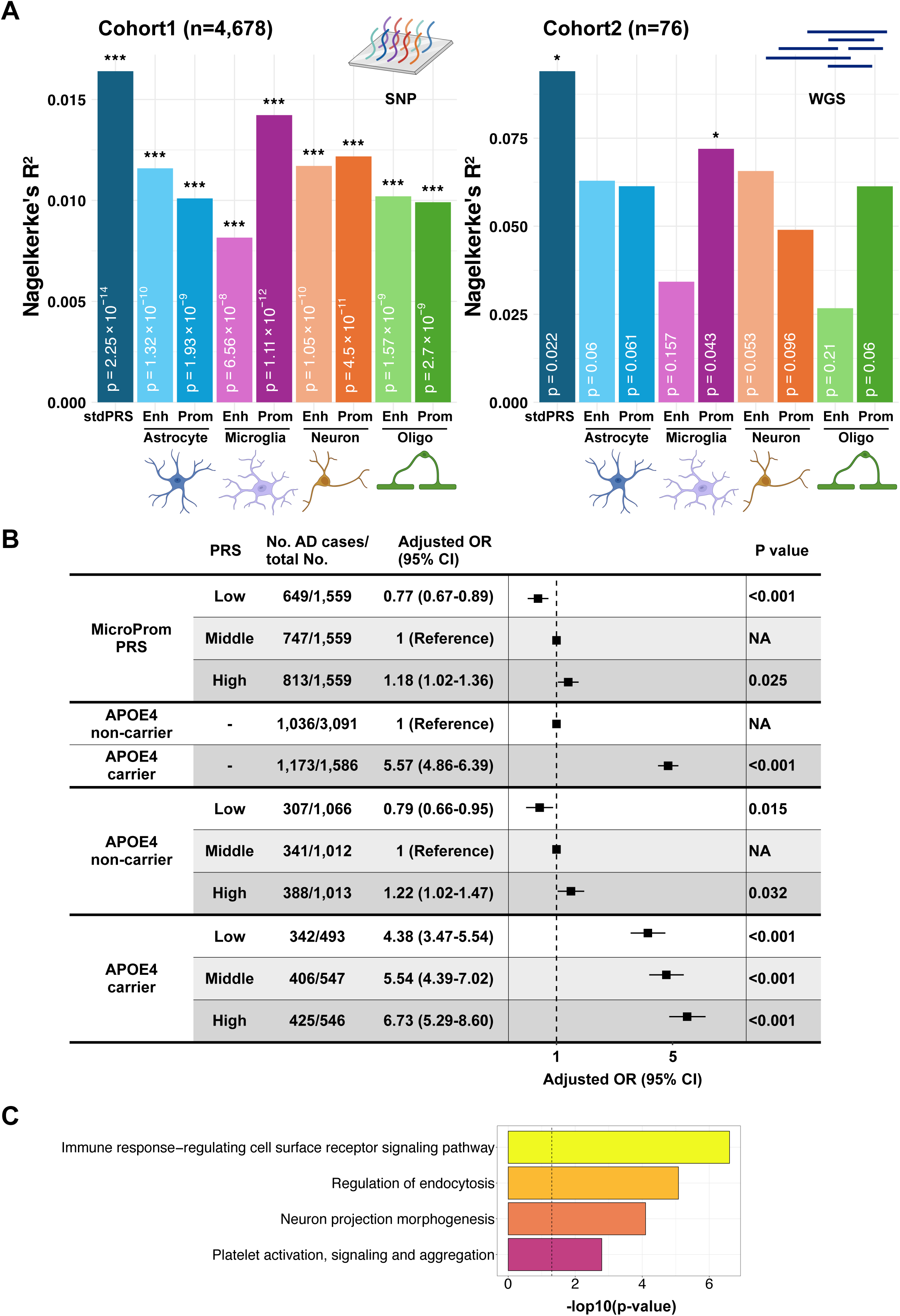
Cell type–specific PRS-based risk assessment for AD. (A) Classification performance of each PRS model across cohorts (*p < 0.05; **p < 0.01; ***p < 1×10⁻³). Created with BioRender.com. (B) Odds ratios for AD risk based on the MicroProm PRS and *APOE* ε4 status. Adjustments were made for the effects of sex and age. P values were calculated using Wald tests. (C) Gene functional enrichment analysis of eGenes included in the MicroProm PRS. Enh, enhancer; Prom, promoter; OR, odds ratio; CI, confidence interval.

Validation in Cohort 2 (PMB cohort) (76 postmortem brains: 39 AD patients and 37 controls; Table 1) revealed that despite the limited sample size, both the standard PRS and MicroProm PRS significantly distinguished AD patients from controls (standard PRS: R² = 0.094, p = 0.022, 78 SNPs; MicroProm PRS: R² = 0.072, p = 0.043, 25 SNPs) (Figures 1A (right panel) and S1, Table S5). Among all cell type–specific models, only the MicroProm PRS was replicated in Cohort 2.

The MicroProm PRS models from both cohorts shared 22 SNPs (88% in Cohort1 and 91.6% in Cohort2). In Cohort 1, the MicroProm PRSs derived from the two cohorts were strongly correlated (Spearman correlation = 0.957, p < 2.2 × 10⁻¹⁶). Using the Cohort 2-derived MicroProm PRS model, we stratified individuals in Cohort 1 into low, middle, and high PRS groups on the basis of tertiles. Compared with the middle PRS group, the low PRS group had a significantly lower risk of AD (OR = 0.77, p = 4.69 × 10⁻⁴), whereas the high PRS group had a significantly increased risk (OR = 1.18, p = 0.025) (Figure 1B). Even when Cohort 1 was stratified into *APOE* ε4 carriers and noncarriers, the MicroProm PRS still contributed to risk stratification (Figure 1B). Given the consistent performance of the MicroProm PRS across both cohorts, we adopted the Cohort 2-derived MicroProm PRS model for all subsequent omics analyses using postmortem brain tissues from Cohort 2.

To explore biological pathways that were potentially influenced by the MicroProm PRS, we examined cis-eQTL data derived from human microglia (see Methods) [40]. This analysis revealed 29 eQTL genes (eGenes) that were regulated by eQTL SNPs included in the MicroProm PRS, many of which were enriched in immune responses and endocytosis pathways (Figure 1C and Table S6).

Collectively, these results define a reproducible microglia-specific PRS model that captures AD genetic risk and implicates immune- and endocytosis-related mechanisms in disease pathogenesis.

### 3.2 Analysis of stratification on the basis of microglia-specific PRS using bulk RNA-seq

We hypothesized that patients with AD and differing polygenic risk scores might present distinct brain pathologies. To test this hypothesis, we performed bulk RNA-seq on prefrontal cortex tissue from Cohort 2 postmortem brains. MicroProm PRS values were calculated for all 100 individuals, and the lowest and highest quartiles were defined as the low and high PRS groups, respectively. AD cases from each group were selected, yielding 6 low-PRS and 10 high-PRS AD patients (Figure 2A).

**Figure 2.**
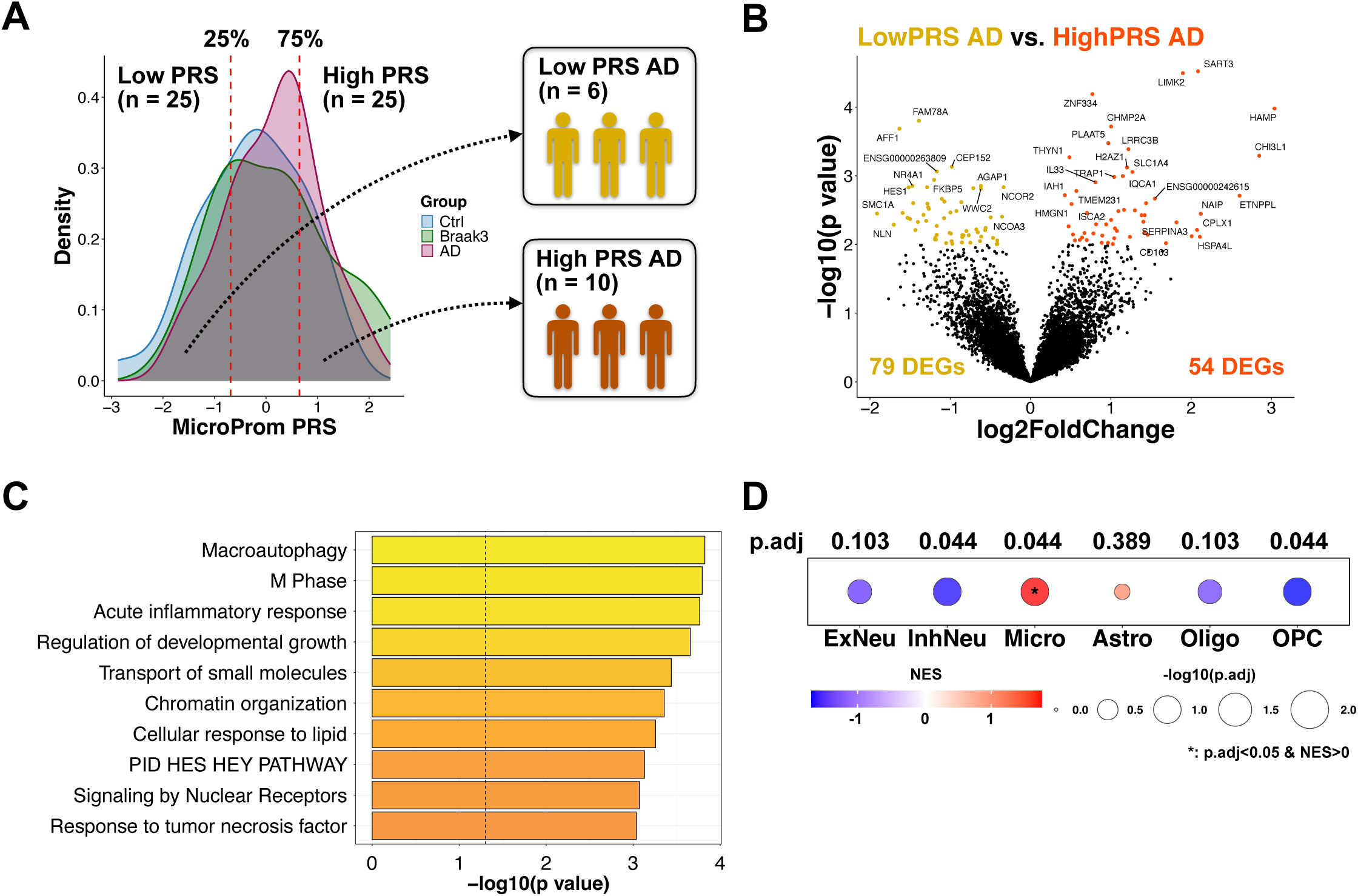
Bulk-brain RNA-seq analysis of the AD subgroups stratified by the MicroProm PRS. (A) Stratification scheme for Cohort 2 (PMB cohort) based on the MicroProm PRS. (B) Volcano plot comparing the low-PRS AD and high-PRS AD groups. DEGs were defined as those for which p < 0.01 and are highlighted. (C) Gene functional enrichment analysis of 112 DEGs identified between the groups. (D) GSEA of DEGs associated with *macroautophagy* or *acute inflammatory response*. Cell-type marker genes were defined using the FindAllMarkers function and used as reference sets, with DEGs as input. NES > 0 indicates enrichment in genes expressed in the corresponding cell type.

RNA-seq revealed 112 differentially expressed genes (DEGs; nominal p < 0.01) between the low- and high-PRS AD groups (Figure 2B), although none survived multiple testing correction. Notably, genes upregulated in the high-PRS AD group included *HAMP*, which encodes hepcidin and is associated with iron accumulation [56], and transmembrane protein 163 (*TMEM163*). Both of these genes are recognized as markers of activated or inflammatory microglia [19–21, 57].

Pathway enrichment analysis revealed that these DEGs were significantly overrepresented in macroautophagy (p = 1.51 × 10⁻⁴) and acute inflammatory response (p = 1.74 × 10⁻⁴) pathways. To evaluate the cell-type specificity of these DEGs, we conducted GSEA using snRNA-seq data obtained from postmortem Japanese brains (analysed in detail in a later section). This analysis revealed that these DEGs were significantly enriched in microglia (NES = 1.75, FDR = 0.044).

These findings suggested that microglial activation states differed between patients with AD stratified by the MicroProm PRS, supporting a mechanistic link between genetic risk and microglial pathology.

### 3.3 Microglial Subcluster Annotation by snRNA-seq

Given that bulk-brain RNA-seq analysis suggested potential differences in microglial states across AD patient subtypes stratified by the microglia polygenic risk score (MicroProm PRS), we performed snRNA-seq on the frontal cortex to directly test whether these PRS-defined subtypes exhibited distinct microglial transcriptional profiles. We selected 15 samples from Cohort 2, comprising three low-PRS AD patients, four high-PRS AD patients, and eight non-AD controls with low MicroProm PRSs, all of which had high-quality RNA (RNA integrity numbers > 7) (Table S3).

After QC, 111,918 nuclei remained for downstream analysis, with a median of 2,067 genes and 7,335 UMIs per nucleus (Figure S2A). Clustering based on canonical cell-type markers revealed eight major brain cell types (Figures 3A, S2B–D). We subsequently isolated the microglial population for reclustering, which yielded eight distinct subclusters (Figure 3B).

**Figure 3.**
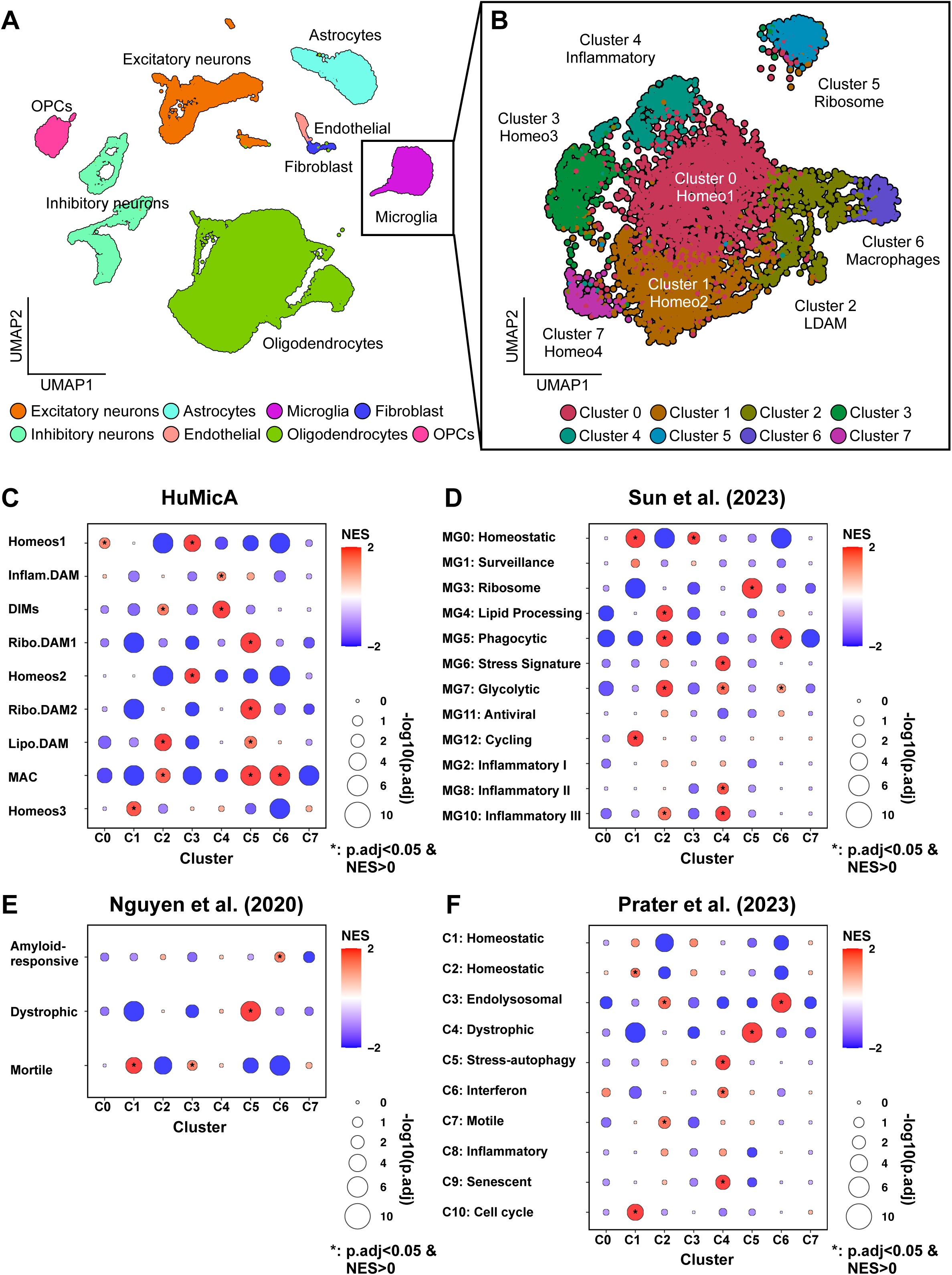
snRNA-seq analysis of the frontal cortex and microglial subcluster annotation. (A) UMAP projection showing annotated cell types. (B) Microglial clustering results. (C–F) GSEA of microglial subclusters. The reference sets consisted of subcluster-specific genes defined using FindAllMarkers. Predefined microglial subcluster marker genes from the HuMicA dataset [20] (C), Sun *et al.* [21] (D), Nguyen *et al.* [42] (E), and Prater *et al.* [43] (F) were used as input sets. NES > 0 indicates upregulation in the subcluster.

Subcluster annotation was guided by two reference datasets: the Sun *et al.* [21] and Human Microglia Atlas (HuMicA) [20] datasets (Figures 3C and 3D). Clusters 0 and 1 expressed homeostatic microglial markers (*CX3CR1* and *P2RY12*) and corresponded to homeostatic clusters in both references. Cluster 3, characterized by *GRID2*, matched the HuMicA Homeos2 cluster and Sun *et al.*’s MG0 subcluster, and was classified as homeostatic. Cluster 7, although not significantly enriched in GSEA, expressed *SYNDIG1* and *ST6GALNAC3*, both of which are found in the Homeos1–3 clusters of the HuMicA dataset and in the MG0 and MG1 subcluster from Sun *et al.* Based on this expression profile, we classified Cluster 7 as a homeostatic microglial subpopulation.

Cluster 2 expressed lipid-associated microglial markers (*GPNMB*, *PTPRG*) and corresponded to the LDAM cluster in HuMicA and the MG4 subcluster in Sun *et al.* Cluster 4 expressed the inflammatory cytokine *SPP1* and was enriched for inflammatory microglial gene sets. Cluster 5 was characterized by high expression of iron-storage genes (*FTL* and *FTH1*) and ribosomal genes, which is consistent with ribosome biogenesis. Cluster 6, with high *F13A1* expression, aligned with macrophage-like populations in HuMicA.

Cluster 5, which was associated with ribosome biogenesis, was distinctly separated from the other clusters in the UMAP space. To further investigate its biological significance, we compared the Cluster 5 gene set to the microglial gene sets from Nguyen *et al.* [42], which classifies microglia into amyloid-responsive, dystrophic, and motile subclusters. The Cluster 5 gene set showed a strong correlation with the dystrophic microglial gene set (Figure 3E). This finding was independently validated in the dataset by Prater *et al.* [43] (Figure 3F). These results suggested that Cluster 5 represented a population of dystrophic microglia that might be structurally and functionally compromised in response to external stress.

In total, we identified eight microglial subclusters: four homeostatic subclusters (Clusters 0, 1, 3, and 7), one lipid-associated subcluster (Cluster 2), one inflammatory subcluster (Cluster 4), one ribosome-associated/dystrophic subcluster (Cluster 5), and one macrophage-like subcluster (Cluster 6).

### 3.4 Analysis of Microglial Stratification Based on Polygenic Risk Using snRNA-seq

To investigate whether microglia-specific genetic risk influences microglial states, we compared the proportions of microglial subclusters— excluding macrophages—across non-AD controls, low-PRS AD patients, and high-PRS AD patients (Figure 4A). Kruskal–Wallis testing revealed a significant reduction in the proportion of homeostatic Cluster 0 in both AD groups relative to the control group, regardless of PRS (p = 0.046). Cluster 2, which displayed LDAM-like features, was relatively increased in the low-PRS AD group, showing a significant increase compared with the combined non-AD control and high-PRS AD groups (p = 0.048, two-sided Wilcoxon rank-sum test). Cluster 5, which was associated with ribosome biogenesis and dystrophic microglia, tended to be more abundant in the high-PRS AD group, although this difference did not reach significance (p = 0.104, two-sided Wilcoxon rank-sum test). We next compared marker gene expression for Clusters 2 and 5. *GPNMB*, *PTPRG*, and *CXCR4*, which defined Cluster 2, were significantly upregulated in the low-PRS AD group (Figure 4B). Ribosomal genes characteristic of Cluster 5 tended to be more highly expressed in the high-PRS AD group, although the difference was not significant (Figure S3A), which was consistent with their subcluster proportions.

**Figure 4.**
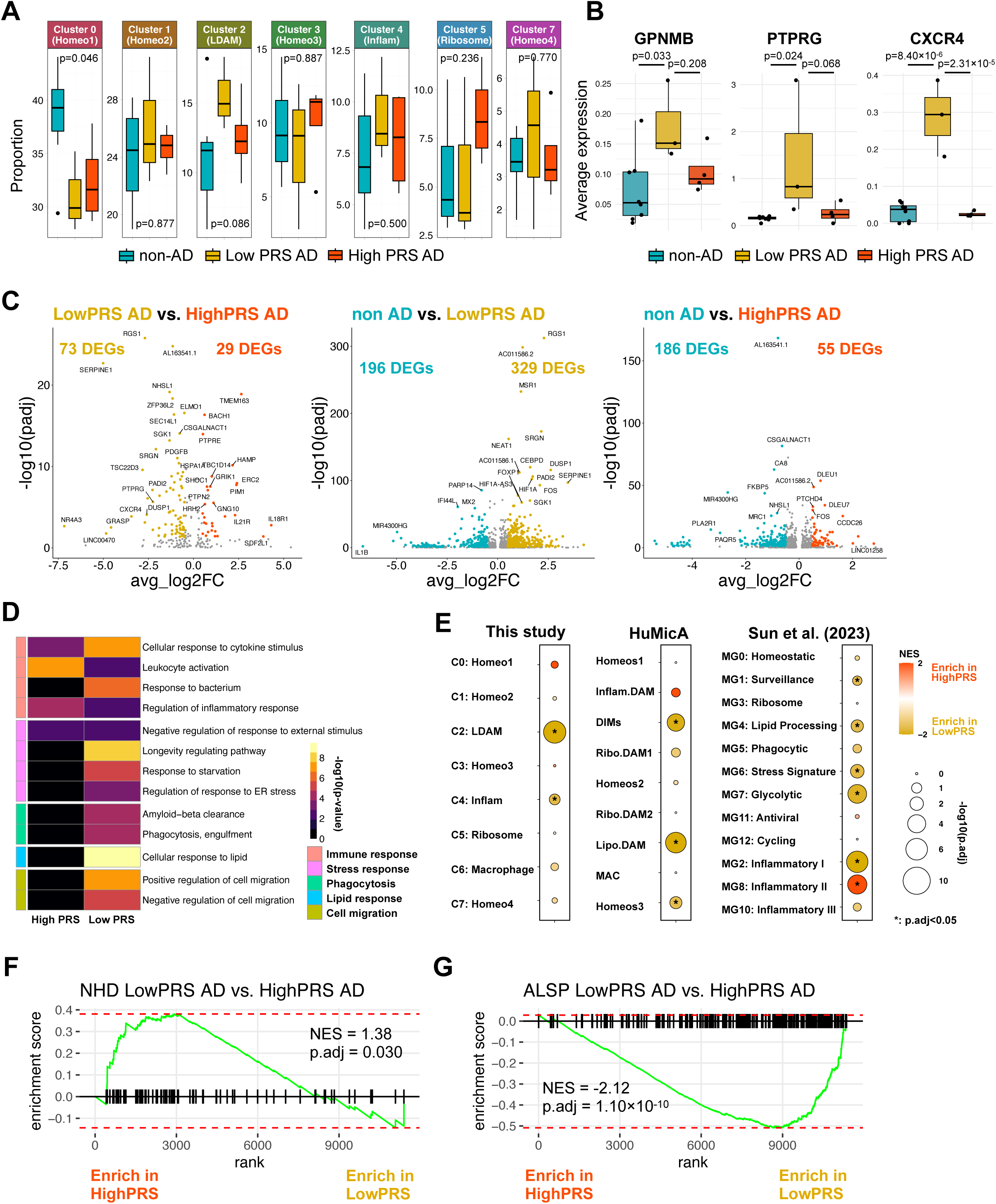
Differences in microglial expression between the low-PRS AD and high-PRS AD groups. (A) Proportions of microglial subclusters. Each proportion was calculated as the proportion of each subcluster relative to the total number of microglia in the sample. P values were calculated by Kruskal–Wallis test. (B) Box plots showing the average expression per sample of LDAM marker genes. P values were calculated by Tukey’s honestly significant difference test. (C) Volcano plot of microglial DEGs. DEGs were defined as genes with adjusted p < 0.05 and |avg_log₂FC| > 0.5. (D) Representative pathways associated with the DEGs between low- and high-PRS ADs. A full list is shown in Figure S3C and Table S7. (E) GSEA of DEGs relative to microglial subclusters. NES > 0 and NES < 0 indicate enrichment in high-PRS AD and enrichment in low-PRS AD, respectively. (F, G) GSEA using signature genes from NHD (F) and ALSP (G).

Differential expression analysis in microglia (adjusted p < 0.05, |log₂FC| > 0.5) revealed 73 genes upregulated in low-PRS AD patients and 29 upregulated in high-PRS AD patients, indicating a 2.5-fold higher DEG count in the low-PRS AD group (left panel in Figure 4C). Notably, the high-PRS AD DEGs included *HAMP* and *TMEM163*, which were both observed in our bulk-brain RNA-seq analysis and associated with inflammatory microglia, whereas the low-PRS AD DEGs included LDAM markers such as *CXCR4* and *PTPRG*. This difference in DEG counts, with a greater number in the low PRS group, persisted under alternative DEG thresholds (Figure S3B). In comparison with non-AD controls, the low-PRS AD group had 329 upregulated genes versus 196 upregulated genes in non-AD controls (1.68-fold) (center panel in Figures 4C, S3B), while the high-PRS AD group had 55 versus 186 upregulated genes in non-AD controls (0.30-fold) (right panel in Figures 4C, S3B). These findings suggested that microglia in low-PRS AD showed greater transcriptional activation, whereas microglia in high-PRS AD exhibited attenuated activation. Gene functional enrichment analysis revealed 386 pathways (Table S7, Figure S3C). Among these pathways, those related to microglial biology included immune and stress responses, which were enriched in both groups, and phagocytosis, lipid response, and cell migration, which were enriched only in the low-PRS AD group (Figure 4D). The genes upregulated in the low-PRS AD group consistently overlapped with those in the LDAM clusters across the three independent datasets (Figure 4E).

Our results indicated that microglia in the high-PRS AD group exhibited attenuated activation, potentially reflecting cumulative genetic risks associated with impaired microglial function. To further investigate the nature of this attenuation, we compared their transcriptomes with those from two prototypical primary microgliopathies with well-defined molecular aetiologies: NHD, caused by loss-of-function mutations in *TREM2* or *TYROBP* that impair microglial phagocytosis, and ALSP, caused by loss-of-function mutations in *CSF1R*, a gene essential for microglial development, proliferation, and survival. Given that the MicroProm PRS captures genetic susceptibility related to endocytosis pathways (Figure 1C), we hypothesized that high-PRS AD might share molecular features with NHD, whereas ALSP, in which LDAM-like microglial activation is also observed as in typical AD [45], would resemble the low-PRS AD group. Using microglial signature genes derived from NHD and ALSP patient brains [44, 45], we assessed the transcriptomic similarity across groups. The NHD signatures showed greater alignment with the high-PRS AD group (NES = 1.38; adjusted p = 0.030; Figure 4F), whereas the ALSP signatures were more strongly associated with the low-PRS AD group (NES = -2.15; p = 1.10 × 10^-10^; Figure 4G). These findings suggested that high-PRS AD microglia adopted impaired phagocytic states reminiscent of NHD pathology, whereas low-PRS AD microglia displayed transcriptional profiles consistent with LDAM-like activation, typical of AD and also observed in ALSP.

### 3.5 Spatial Transcriptomic Profiling Based on Microglial PRS Stratification

To validate and spatially resolve the PRS-dependent microglial states observed via snRNA-seq, we performed spatial transcriptomic analysis using the GeoMx DSP–WTA platform on eight postmortem brain samples (three low-PRS AD, four high-PRS AD, and one non-AD control). All the samples were also included in the preceding snRNA-seq analysis.

ROIs were defined by immunohistochemistry for IBA1 (microglia), GFAP (astrocytes), and Aβ plaques, enabling the identification of glial cells and plaque pathology. Within each ROI, AOIs enriched in microglia were selected while minimizing astrocytic signal contamination. AOIs were categorized as follows: on-plaque microglia (IBA1⁺Aβ⁺GFAP⁻) within Aβ deposits and periplaque microglia (IBA1⁺Aβ⁻GFAP⁻) located adjacent to Aβ plaques (Figure 5A). Following stringent QC (see Methods), at least two AOIs were obtained from two individuals per group for both conditions. As no AOIs from the non-AD control passed QC, we focused subsequent analyses on the two AD groups.

**Figure 5.**
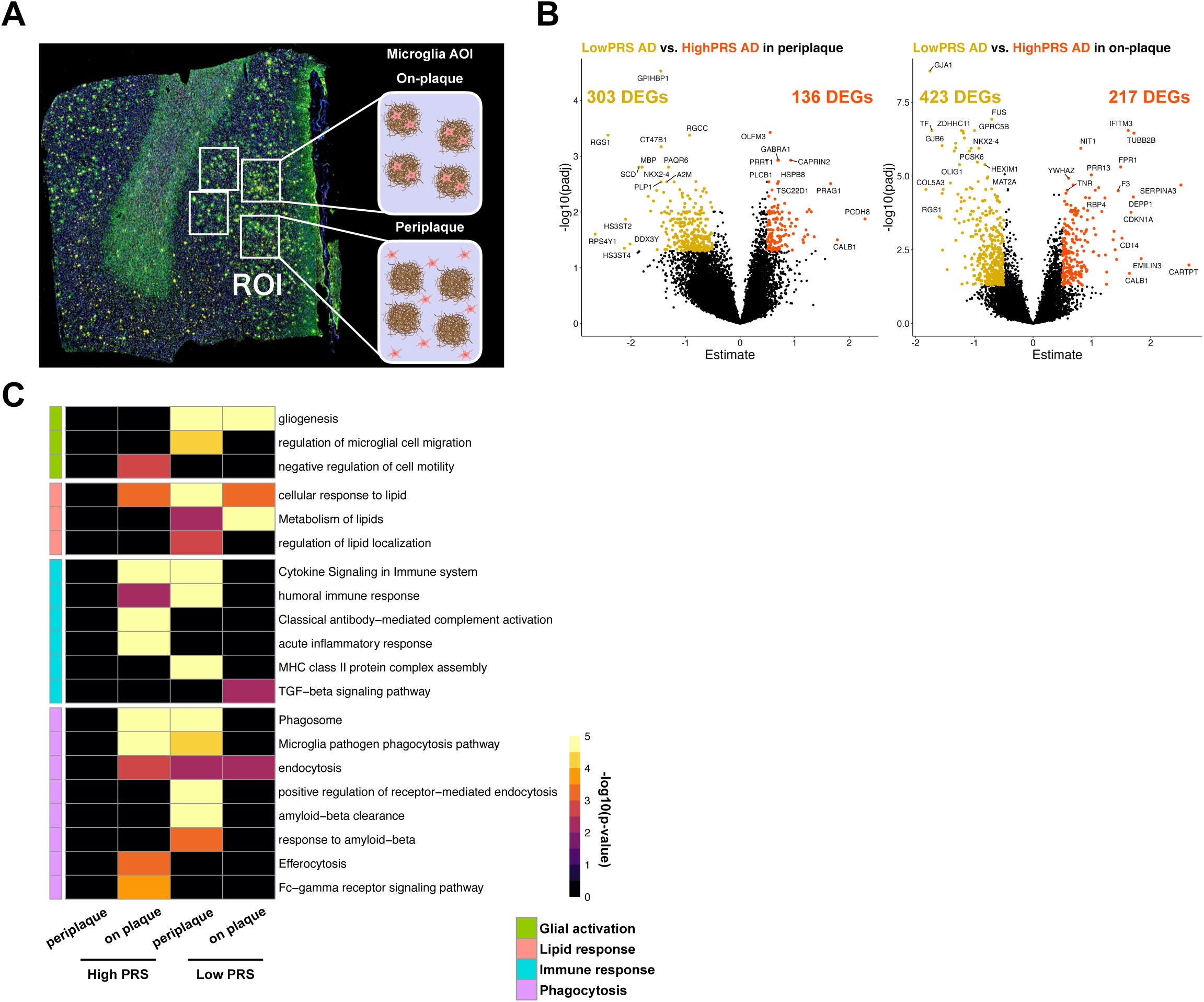
Spatial transcriptomic analysis of microglia in the low-PRS AD and high-PRS AD groups. (A) Schematic of periplaque and on-plaque regions. (B) Volcano plots of DEGs in periplaque and on-plaque microglia. DEGs were defined as genes with adjusted p < 0.05 and |Estimate| > 0.5. (C) Representative pathways associated with DEGs from each region. A full list is shown in Table S8.

Differential gene expression analysis between the high- and low-PRS AD groups, using linear mixed-effects models, defined DEGs as those with adjusted p value < 0.05 and |log₂FC| > 0.5. In periplaque microglia, 303 genes were upregulated in the low-PRS AD group, and 136 genes were upregulated in the high-PRS AD group (left panel in Figure 5B). Similarly, in on-plaque microglia, 423 and 217 genes were upregulated in the low-PRS AD group and high-PRS AD group, respectively (right panel in Figure 5B). Thus, the low PRS AD group consistently presented more upregulated genes, with 2.23-fold and 1.95-fold higher counts in the periplaque and on-plaque conditions, respectively. This pattern persisted under alternative thresholds (Figure S4A) and paralleled the snRNA-seq findings, supporting greater transcriptional activation in low-PRS AD microglia.

We next performed gene functional enrichment analysis of the upregulated genes in each group. Across all the conditions, 1,637 pathways were significantly enriched in at least one group (Table S8). From these, we highlight representative pathways in four categories of biological processes with established relevance to microglial function—glial activation, lipid metabolism, immune response, and phagocytosis (Figure 5C). Among the pathways related to glial activation, “gliogenesis” was significantly enriched only in the low-PRS AD group, suggesting enhanced glial proliferation in response to Aβ. In periplaque microglia, the low-PRS AD group showed enrichment of “regulation of microglial cell migration”, whereas the high-PRS AD group showed “negative regulation of cell motility” in on-plaque microglia, indicating differential microglial motility between the groups. Lipid metabolism–related pathways were prominently upregulated in low-PRS microglia, which is consistent with the presence of LDAM states. While both groups showed enrichment of immune response pathways, the specific pathways differed between the groups. The high PRS AD group showed specific enrichment of the “acute inflammatory response” in on-plaque microglia, whereas the low PRS AD group was characterized by upregulation of “MHC class II protein complex assembly” and activation of the “TGF-beta signalling pathway”. Phagocytosis-related pathways, including “phagosome formation” and “endocytosis”, were enriched in both groups, but there were differences in downstream degradation activity. Low PRS AD periplaque microglia showed “Aβ clearance”, whereas high PRS AD on-plaque microglia showed “Fc-gamma receptor signalling pathway” and “efferocytosis”, indicating apoptotic cell removal.

Notably, genes that were upregulated in periplaque microglia in the high-PRS AD group were not enriched in the above described pathways. Independent pathway analysis of these genes revealed enrichment in “neuronal system” and “synaptic transmission” pathways (Figure S4C), potentially reflecting neuronal engulfment processes linked to efferocytotic activity.

Collectively, these results indicated that low-PRS AD microglia exhibited stronger transcriptional activation, particularly in pathways related to Aβ clearance and lipid metabolism, which was consistent with the snRNA-seq results and suggested that the phagocytic response was more active in low-PRS AD microglia than in high-PRS AD microglia.

### 3.6 Differences in phagocytic activity by PRS stratification

We focused on the phagocytic pathway identified in the pathway analysis and sought to clarify how phagocytosis and its downstream processes differed between AD groups stratified by PRS. On the basis of the expression patterns of 75 DEGs included in the *Phagocytosis* category shown in Figure 5C, these genes were classified into six groups (Figures 6A and S5A).

**Figure 6.**
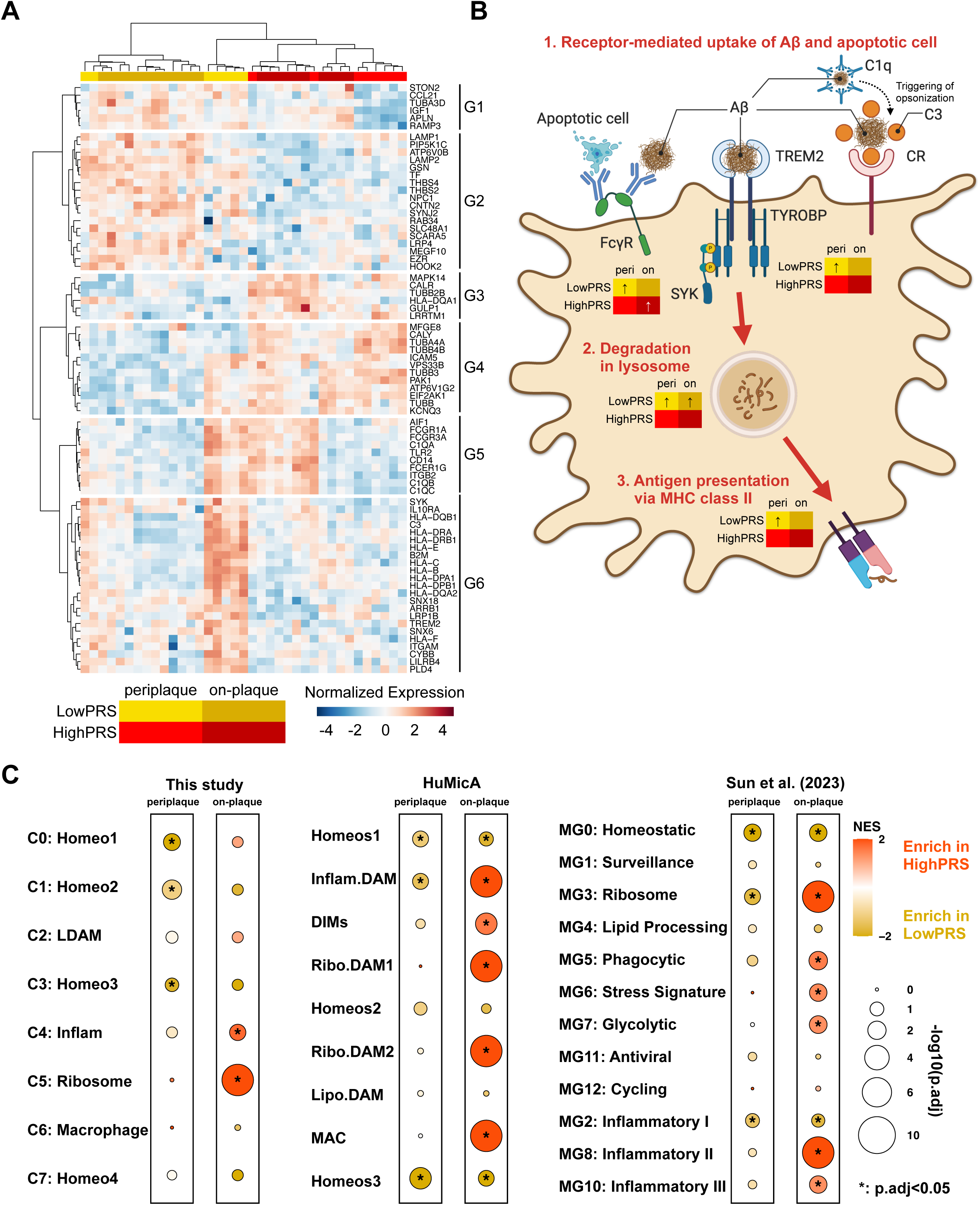
Comparative expression of genes related to phagocytosis, degradation, and antigen presentation in microglia. (A) Heatmap of DEGs related to phagocytosis. (B) Schematic of phagocytosis– degradation–antigen presentation pathways. Created with BioRender.com. (C) GSEA of DEGs relative to microglial subclusters. NES > 0 and NES < 0 indicate enrichment in high-PRS AD and in low-PRS AD, respectively.

Group 2 included lysosomal genes (*LAMP1* and *LAMP2*), which were upregulated in both periplaque and on-plaque microglia in the low-PRS AD group, consistent with the increased Aβ degradation. Group 5 included Fc receptors (*FCGR1A*, *FCGR3A*, and *FCER1G*) and the C1q complex (*C1QA/B/C*), which is a key component of the classical complement pathway and promotes complement-mediated opsonization. This group was enriched in periplaque microglia in the low-PRS AD group and in on-plaque microglia in the high-PRS AD group. Group 6 was specifically upregulated in the periplaque microglia of the low-PRS AD group and contained genes involved in Aβ phagocytosis via the *TREM2–SYK* pathway and the classical pathway (*C3* and *ITGAM*), along with *HLA* genes, indicating the activation of antigen presentation. In addition, *TYROBP*, which is an adaptor of TREM2, was also upregulated in periplaque microglia in the low-PRS AD group, although this increase did not reach statistical significance (Figure S5A).

These findings indicated that in the high-PRS AD group, phagocytosis might be engaged primarily through Fc receptor pathways in on-plaque microglia, whereas downstream processes such as lysosomal degradation and antigen presentation were less activated. In contrast, periplaque microglia in the low-PRS AD group exhibited coordinated upregulation of pathways involved in phagocytosis, degradation, and antigen presentation (Figure 6B).

Finally, we examined the microglial subclusters associated with the DEGs between low- and high-PRS ADs in each spatial location. Genes upregulated in on-plaque microglia in the high-PRS AD group overlapped significantly with ribosome-associated microglia (dystrophic microglia) markers across multiple reference datasets (Figure 6C). These findings were consistent with our snRNA-seq results showing upregulation of ribosome biogenesis genes in the high-PRS AD group (Figure S3A) and were further supported by the elevated expression of ribosomal genes in spatial transcriptomic data from high-PRS on-plaque microglia (Figure S5B). On the basis of these findings, we concluded that microglia in the high-PRS AD group presented dysfunctional phagocytosis and preferentially exhibited a dystrophic state.

### 3.7 Microglia-Specific Signatures and Aβ Responses in the Low-PRS AD Group

To explore differences in microglial responses to Aβ according to genetic background, we sought to identify microglia-specific signature genes for the low- and high-PRS AD groups. We first screened for genes with consistently upregulated expression in both the snRNA-seq and spatial transcriptomics analyses. No overlapping genes were detected in the high-PRS AD group across the two modalities. In contrast, the expression of six genes (*CXCR4*, *RGS1*, *DUSP1*, *SGK1*, *GLDN*, and *ZFP36L2*) was consistently upregulated in the low-PRS AD group (Figure 7A).

**Figure 7.**
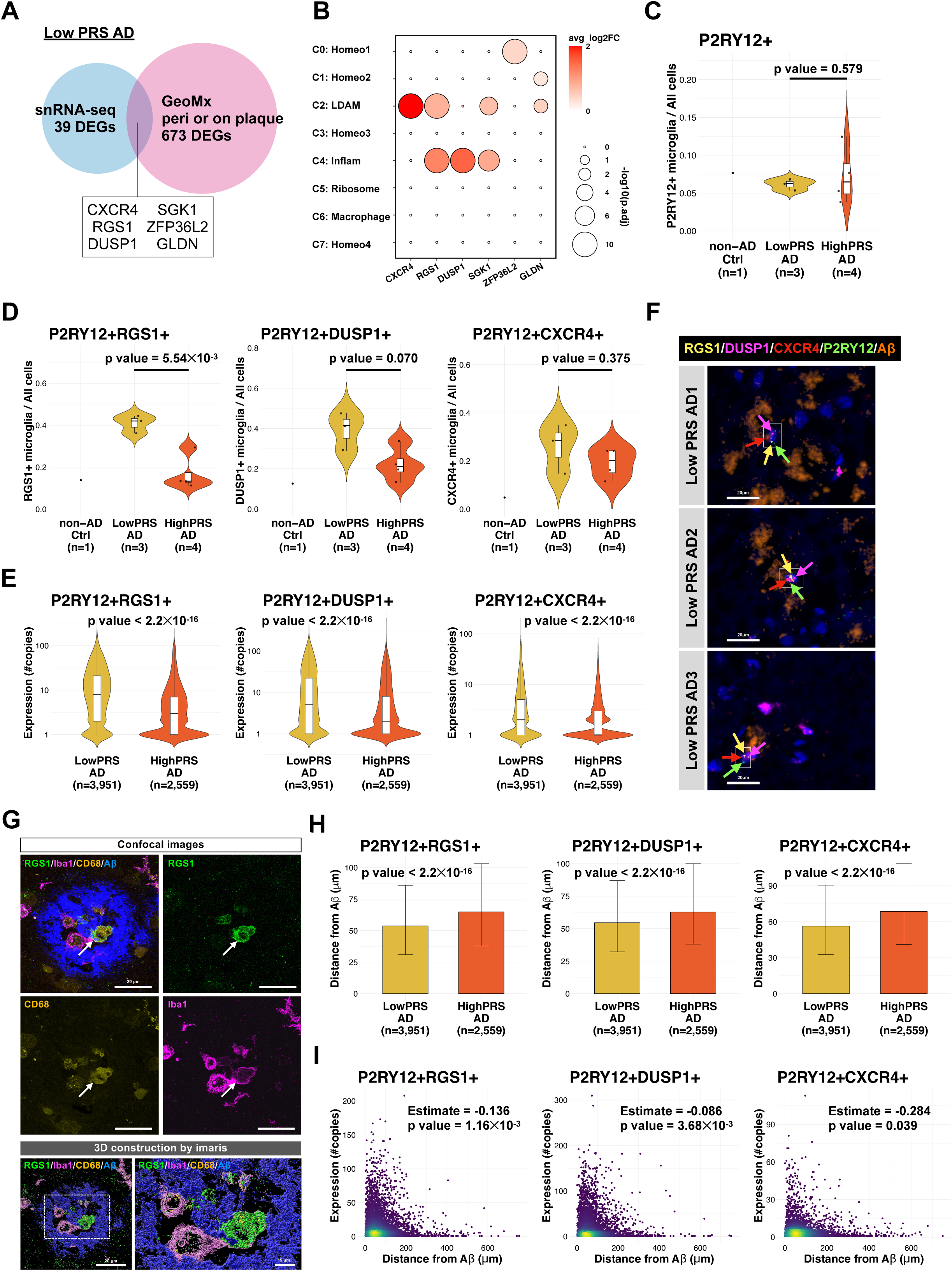
Characterization of microglia expressing low-PRS AD signature genes. (A) Overlap of genes upregulated in low-PRS AD identified by snRNA-seq and spatial transcriptomics. (B) Distribution of signature gene expression across microglial subclusters. (C) Proportions of microglial cells. Each proportion was calculated as the fraction of P2RY12⁺ cells relative to all cells identified by the HALO software. The non-AD control sample was excluded from a statistical test because n = 1. (D) Proportion of P2RY12⁺ microglia expressing each signature gene of low-PRS AD. (E) Expression levels of P2RY12⁺ in microglia expressing each signature gene. P values were calculated by a two-sided Wilcoxon rank-sum test. (F) Immunohistochemistry for Aβ (orange) and RNAscope in situ hybridization for P2RY12 (green), RGS1 (yellow), DUSP1 (magenta), and CXCR4 (red), counterstained with DAPI (blue). Representative images correspond to postmortem frontal cortex tissue sections from three low-PRS AD patients. The arrows indicate single marker-positive dots. The rectangles indicate cells that were classified by the HALO software. Scale bars: 20 μm. (G) Representative confocal images of cerebral cortices from a low-PRS AD patient immunostained with antibodies against RGS1 (green), microglial cell marker Iba1 (magenta) and phagocytic microglia marker CD68 (yellow) together with Aβ antibody (blue). Scale bars: 20 μm. The lower panels show Imaris 3D reconstructed images. Lower right panel shows high-magnification views of the white dotted square in the left panel. Scale bars: 5 μm (high-magnification views). (H) The shortest distance from Aβ plaques for P2RY12⁺ microglia expressing each signature gene. The plots indicate the median ± IQR. P values were calculated by a two-sided Wilcoxon rank-sum test. (I) Correlations between signature gene expression and Aβ distance. Slopes and p values were calculated by linear regression and the Wald test, respectively.

Next, we evaluated the associations of these six genes with microglial subclusters (Figure 7B). *CXCR4* was a marker of LDAM, *RGS1* and *SGK1* were expressed in both LDAM and inflammatory microglia, and *DUSP1* was particularly enriched in inflammatory microglia. *GLDN* and *ZFP36L2* appeared to be associated with homeostatic microglia. On the basis of these patterns, we selected *CXCR4* (LDAM marker), *DUSP1* (inflammatory microglia marker), and *RGS1* (more distinct subcluster-specific expression than *SGK1*) as representative signature genes for low-PRS-AD microglia.

To determine whether microglia expressing these three signature genes were more abundant in the low-PRS AD group, we performed immunohistochemistry and RNAscope in situ hybridization using the same eight postmortem brain samples analysed by spatial transcriptomics (three low-PRS AD, four high-PRS AD, and one cognitively normal control). Fluorescent Aβ immunostaining was combined with RNAscope targeting *CXCR4*, *RGS1*, and *DUSP1*, along with the microglial marker *P2RY12*.

Image analysis using HALO software revealed an average of 53,777 cells per sample, with *P2RY12*-positive cells (*i.e.*, microglia) comprising 6.9% on average. This proportion closely matched the microglial fraction (6.3%) from our snRNA-seq analysis, and no significant difference was observed between groups (p = 0.579; Figure 7C), indicating comparable microglial detectability across the samples.

Next, we evaluated the proportion of *P2RY12*-positive microglia that also expressed each signature gene of the low-PRS AD. *P2RY12*-positive cells expressing the signature genes tended to increase in the low-PRS AD group, with the number of *RGS1*-positive microglia significantly increasing (p = 5.54 × 10⁻³; Figure 7D). In microglia expressing each signature gene, the expression of these genes was readily detectable in low-PRS AD samples (Figure 7E). Immunohistochemical and RNAscope analyses confirmed the presence of microglia expressing signature genes in tissue sections from low-PRS AD patients (Figures 7F and S6). We confirmed the presence of RGS1-expressing microglia by immunostaining and observed their co-localization with the phagocytic marker CD68 (Figure 7G).

Given our spatial transcriptomics findings that Aβ clearance pathways were activated in low-PRS-AD microglia (Figure 5C), we hypothesized that these cells might preferentially cluster near Aβ deposits. To test this hypothesis, we measured the shortest distance between microglia expressing each signature gene and Aβ deposits and compared the distances between groups. With respect to all three signature genes, the microglia in the low-PRS AD group tended to be located closer to Aβ (Figure 7H), suggesting enhanced Aβ-associated clustering.

Finally, to assess whether the expression of the signature genes was induced by Aβ, we examined correlations between the gene expression intensity and the distance to Aβ deposits across all AD samples. For each gene, the expression intensity was significantly negatively correlated with the distance from Aβ (Figure 7I), indicating that the expression of these genes was associated with the closer proximity to Aβ.

## 4. Discussion

In this study, we constructed cell type-specific PRSs based on enhancer and promoter regions active in major brain cell types, including neurons, microglia, astrocytes, and oligodendrocytes. An evaluation of eight cell type-specific PRS models across two independent Japanese cohorts revealed that the PRS derived from SNPs enriched in microglial promoter regions (MicroProm PRS) consistently demonstrated the highest discriminative performance for AD. This finding was consistent with recent AD GWASs, which have identified disease-associated loci predominantly in the vicinity of microglia-related genes [15].

In the larger discovery cohort (Cohort 1), optimal predictive performance was achieved using relatively few SNPs (≤78) across all the models, including the standard PRS. This pattern aligned with prior AD PRS studies, suggesting that AD might follow an oligogenic rather than a polygenic architecture, with risk driven by a limited number of moderate-effect SNPs [3, 6, 58, 59]. In Cohort 2, the number of SNPs included in each model varied, likely reflecting limitations in sample size (Table S2).

Similarly, a previous study revealed that microglia-based PRSs exhibit strong discriminative power, second only to the conventional PRS that incorporates all the SNPs [12]. Here, the MicroProm PRS demonstrated high reproducibility when constructed independently in both cohorts, underscoring its robustness and generalizability. These findings further supported the involvement of microglial genetic risk in AD pathogenesis, even in non-European populations.

Using the MicroProm PRS, we stratified AD patients into high- and low-PRS groups and investigated the molecular differences between them. snRNA-seq and spatial transcriptomic analyses revealed enrichment of LDAM in the low-PRS AD group, whereas the high-PRS AD group was characterized by ribosome-associated microglia with features of dystrophic microglia.

LDAM has recently been recognized as a prominent microglial state in AD brains [19–21]. Their upregulation of lipid metabolism- and phagocytosis-related genes suggests active engagement in Aβ clearance. While intracellular lipid droplet accumulation may support energy storage and mitigate ER stress and lipotoxicity, excessive lipid loading can provoke inflammation and damage neighbouring astrocytes and neurons [60]. Thus, the presence of LDAM may reflect a compensatory adaptation to stress, but its overrepresentation could indicate pathological overload.

In contrast, although ribosome-associated microglia have been identified in multiple studies [19–21], their functional characteristics remain poorly understood. Our findings demonstrated that the transcriptomic profile of ribosome-associated microglia closely resembled that of dystrophic microglia, suggesting that this state might reflect a degeneration-prone, stressed microglial phenotype induced by ageing and Aβ accumulation. Dystrophic microglia have long been known to express ferritin, a key marker that is also upregulated in ribosome-associated microglia [61]. Therefore, our observation was not entirely unexpected. Nevertheless, our study provides a valuable link between ribosome-associated microglia frequently detected in single-cell AD datasets and their underlying pathological correlates.

Comparative analyses with microglial genetic disorders revealed that the expression profiles of the high-PRS AD group were similar to those observed in NHD, whereas those of the low-PRS AD group resembled those of ALSP. Loss-of-function mutations in *TREM2*, the causative gene for NHD, impair microglial phagocytosis, and patients with AD carrying *TREM2* variants (e.g., R47H and R62H) exhibit attenuated microglial responses [62]. In Trem2-deficient mouse models, impaired microglial clustering around Aβ plaques exacerbates pathology [63, 64], which was consistent with our observations in the high-PRS AD group.

In contrast, *CSF1R*, the causative gene for ALSP, is essential for microglial homeostasis. Its dysfunction may irreversibly drive microglial transition from homeostatic to activated states. Postmortem analyses of ALSP brains have reported the upregulation of LDAM-related genes such as *CXCR4* and overlaps in gene signatures with AD and multiple sclerosis [45]. These findings are consistent with the observed increase in the LDAM in the low-PRS AD group.

To identify gene signatures characterizing the low-PRS AD group, we focused on six genes that were consistently upregulated according to both the snRNA-seq and spatial transcriptomic analyses. Among them, we focused on *CXCR4*, *RGS1*, and *DUSP1*. *CXCR4* encodes a chemokine receptor involved in microglial migration, aggregation around Aβ plaques, and modulation of inflammatory and anti-inflammatory microglial phenotypes [65–68]. *RGS1*, a negative regulator of *CXCR4* signalling, may promote localized microglial responses and enhance phagocytic activity. Individuals carrying an AD protective allele (rs1921622) exhibit elevated *RGS1* expression and enhanced Aβ phagocytosis by microglia [57]. *DUSP1*, a phosphatase that suppresses the MAPK pathway, has been reported to exert anti-inflammatory and neuroprotective effects in microglia [69–72]. While *RGS1* and *DUSP1* both act to suppress microglial motility, *CXCR4* may represent a distinct population of migratory microglia. We speculate that *CXCR4*-positive microglia actively detect and migrate towards Aβ, whereas *RGS1*- and *DUSP1*-positive microglia may remain localized and focus on phagocytosis. However, we cannot exclude the possibility that these latter cells represent nonmigratory, dysfunctional microglia, warranting further investigation.

Our study demonstrated that stratification of AD patients using the MicroProm PRS captures distinct microglial states and differential responses to Aβ. These findings enhance our understanding of genetically driven molecular heterogeneity in AD and may provide a foundation for precision medicine approaches. For instance, in low-PRS AD, suppressing excessive microglial responses may be beneficial for disease modification, whereas in high-PRS AD, therapeutic strategies aimed at enhancing impaired microglial function may be more effective. Furthermore, our results highlight the potential of PRS not only as a risk predictor but also as a tool for disease subtyping.

Nonetheless, this study has several limitations. First, the number of samples analysed was limited. Larger cohorts and cost-effective multiomics technologies are required for future validation. Second, the GWAS summary statistics used to construct the PRS were derived from populations with different ancestral backgrounds. Although ancestry-matched GWAS data would be ideal, large-scale Japanese GWASs remain lacking. Future studies using ancestry-concordant data may improve PRS accuracy. That said, previous studies have demonstrated good transferability of European-based PRSs to non-European populations, including Japanese [8], lending support to the validity of our findings. Third, functional validation of the identified microglial subtypes in vitro and in vivo will be necessary to confirm their roles in Aβ clearance and disease progression.

## AUTHOR CONTRIBUTIONS

MK: Study design, analysis and interpretation of data, and drafting of the manuscript. AM, NH, MH, KO, SN: Genotyping analysis, Sequencing analysis, interpretation of data, and revision of the manuscript. MM, YSaito, SM: Provision of autopsy brains and revision of the manuscript. YH, YSakakibara, MS, KMI: Immunohistochemistry, interpretation of data and revision of the manuscript. TI: Study design, interpretation of data, and drafting of the manuscript. All the authors have read and approved the final manuscript.

## Acknowledgements

We thank all the donors and facility staff for providing autopsy brains and all the participants and staff of the J-ADNI. The J-ADNI was supported by the following funding sources: the Translational Research Promotion Project from the New Energy and Industrial Technology Development Organization of Japan; Research on Dementia, Health Labor Sciences Research Grant; the Life Science Database Integration Project of Japan Science and Technology Agency; the Research Association of Biotechnology (Astellas Pharma Inc., Bristol-Myers Squibb, Daiichi-Sankyo, Eisai, Eli Lilly and Company, Merck-Banyu, Mitsubishi Tanabe Pharma, Pfizer Inc., Shionogi & Co., Ltd., Sumitomo Dainippon, and Takeda Pharmaceutical Company), Japan; and a grant from an anonymous foundation. The reference genome data used for this research were originally obtained from participants in the Tailor-made Medical Treatment Program (BioBank Japan: BBJ), led by Prof. Michiaki Kubo; these data are available at the website of the NBDC Human Database/the Japan Science and Technology Agency (JST). We thank Dr. Atsushi Watanabe at National Center for Geriatrics and Gerontology for providing technical support. We also thank Drs. Shigenori Nonaka and Hiroshi Yoke for Imaris analysis, supported by Joint Research of the Exploratory Research Center on Life and Living Systems (ExCELLS), ExCELLS program No, 23EXC354.

## Ethics approval and consent to participate

The study protocol was reviewed and approved by the central institutional ethics committee at Niigata University, which served as the single review board for all participating institutions, including Niigata University, Tokyo Metropolitan Institute for Geriatrics and Gerontology, and National Center for Geriatrics and Gerontology (approval numbers G2018-0034 and 2024-0111). For J-ADNI, ethics approval was obtained from the review boards of the participating institutions. Informed consent was obtained from every participant prior to enrolment.

## Data availability

All data produced will be available online at NBDC (registration application in progress).

## Figure legends

Table S1. Summary of the participants in Cohort1.

Table S2. Distribution of regulatory regions in each cell type.

Table S3. ROI/AOIs meeting the inclusion criteria.

Table S4. Performance of each PRS model.

Table S5. eGenes regulated by eQTL SNPs included in the MicroProm PRS.

Table S6. Enriched pathways associated with DEGs between the low-PRS AD and high-PRS AD groups.

Table S7. Functional pathways enriched in microglia from the low-PRS AD and high-PRS AD groups by spatial location.

---

**Figure S1.**
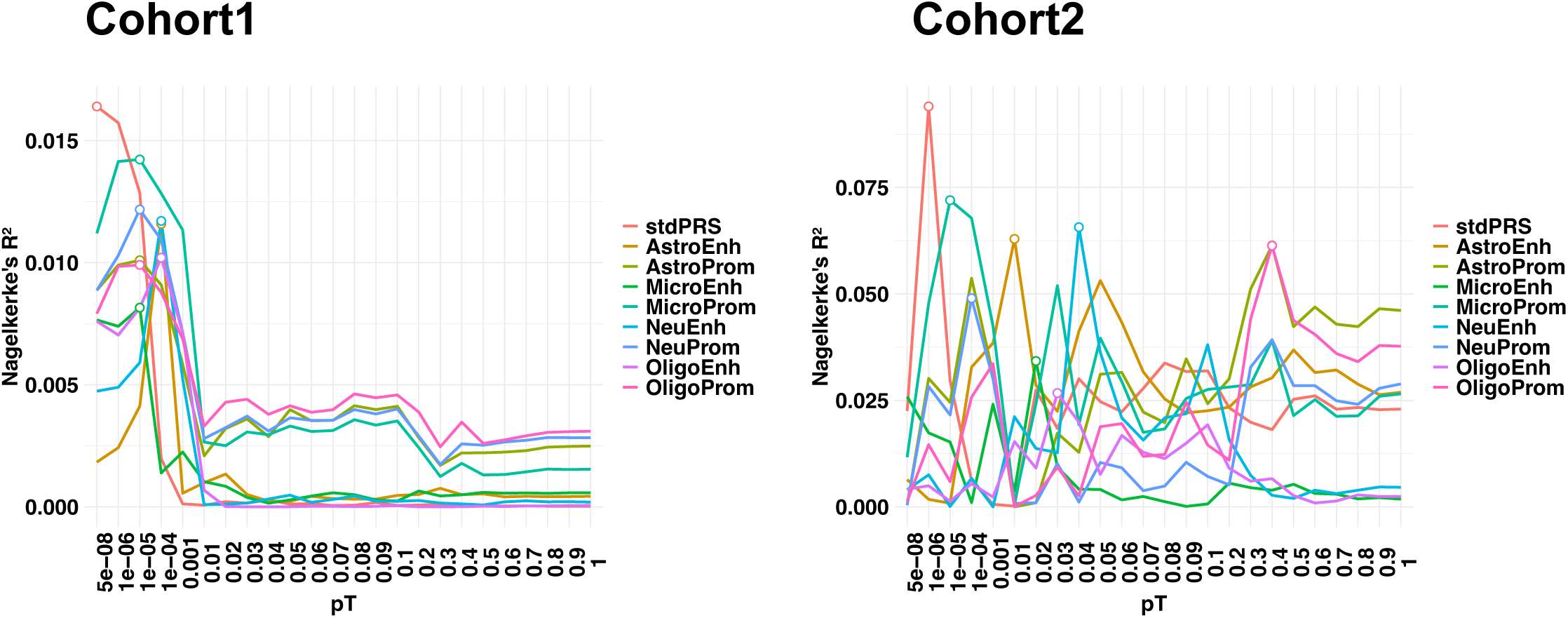
Nagelkerke’s R² across pT thresholds. Open circles indicate the pT values with the best model fit.

**Figure S2.**
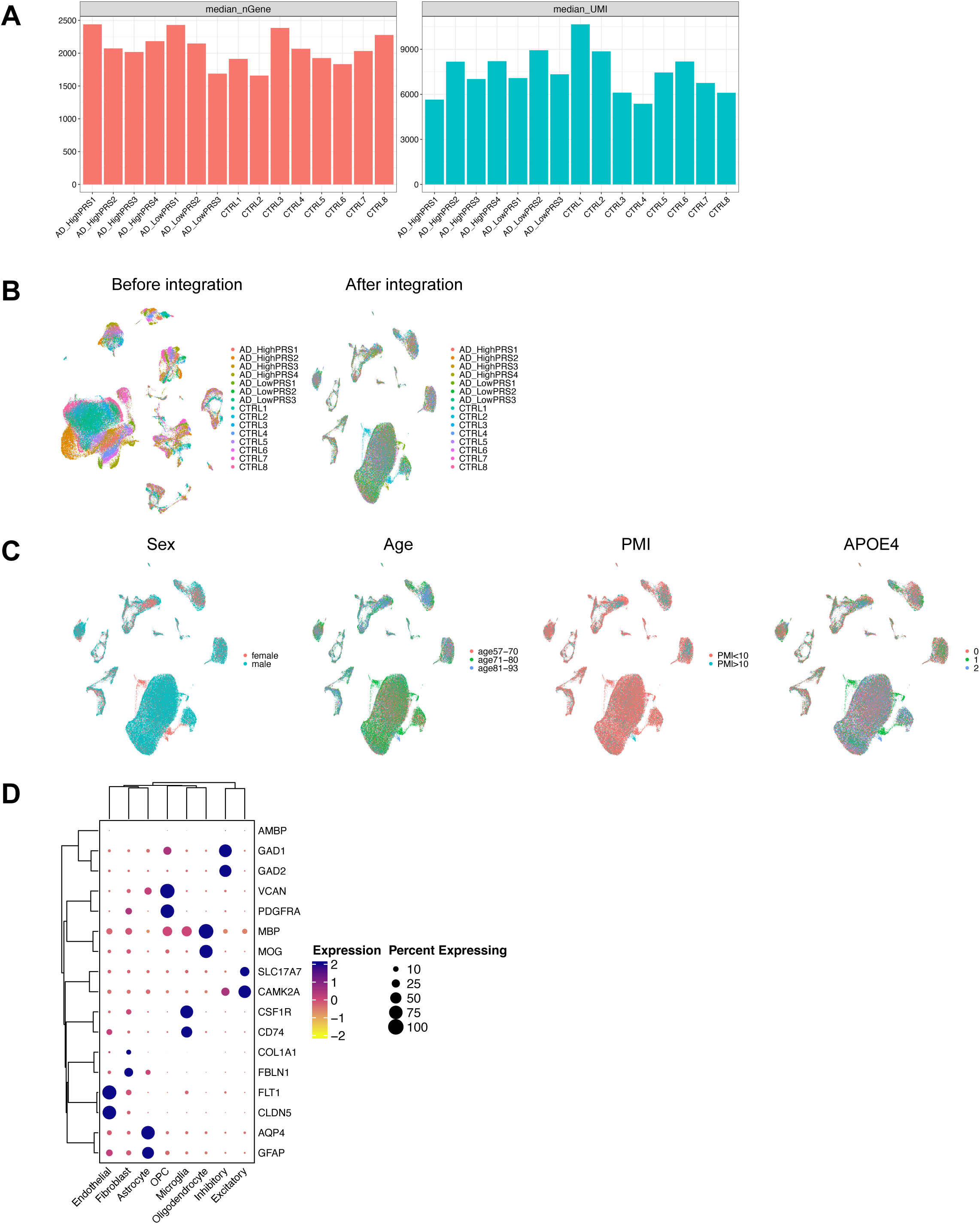
Quality control and characteristics of the snRNA-seq data. (A) Bar plots showing median gene counts and UMI counts per sample. (B) UMAP projections before and after Harmony batch correction. (C) UMAP projections coloured by sample attributes. (D) Dot plots of canonical cell type marker expression.

**Figure S3.**
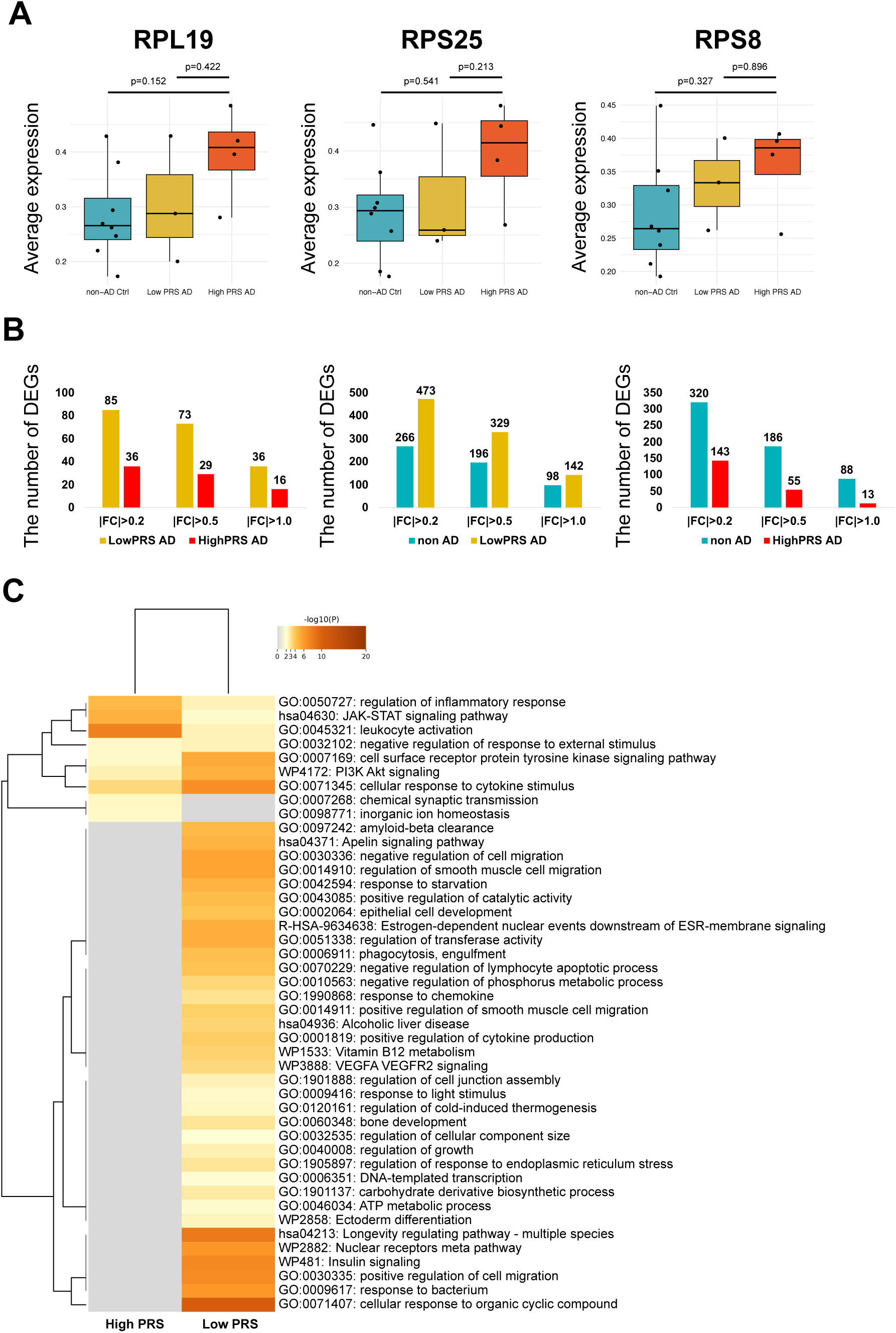
Microglial gene expression analysis in low-PRS AD and high-PRS AD. (A) Box plots showing the average expression of ribosomal genes per sample. (B) Bar plots of DEG counts based on three statistical criteria. (C) Pathways associated with DEGs. Pathways indicate parent terms. A full list is shown in Table S7.

**Figure S4.**
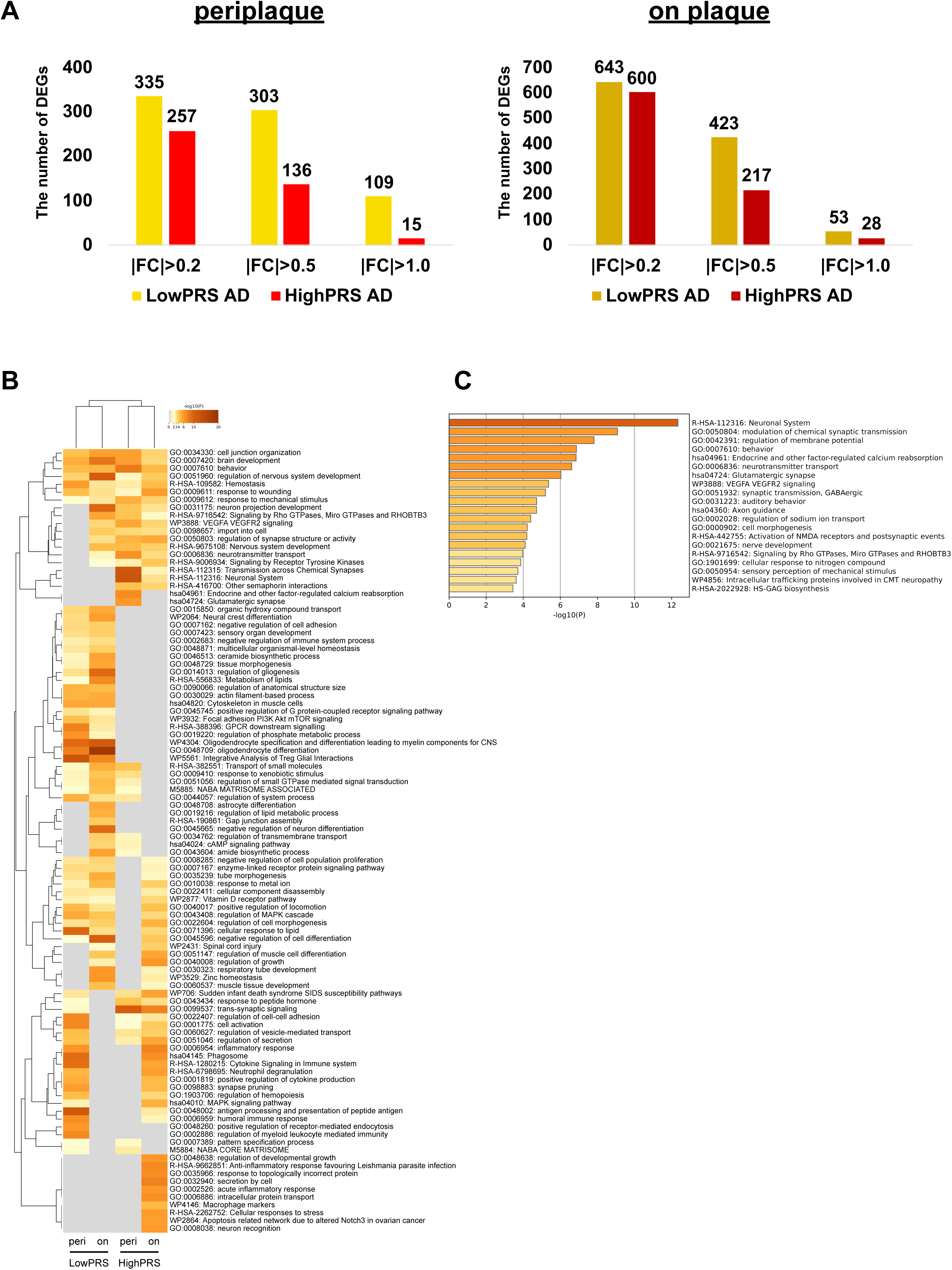
Comparisons of microglial expression through spatial transcriptomics. (A) Bar plots of DEG counts based on three statistical criteria in periplaque and on-plaque regions. (B) Pathways associated with DEGs. Pathways indicate parent terms. A full list is shown in Table S8. (C) Gene functional enrichment analysis of genes upregulated in high-PRS AD in periplaque microglia.

**Figure S5.**
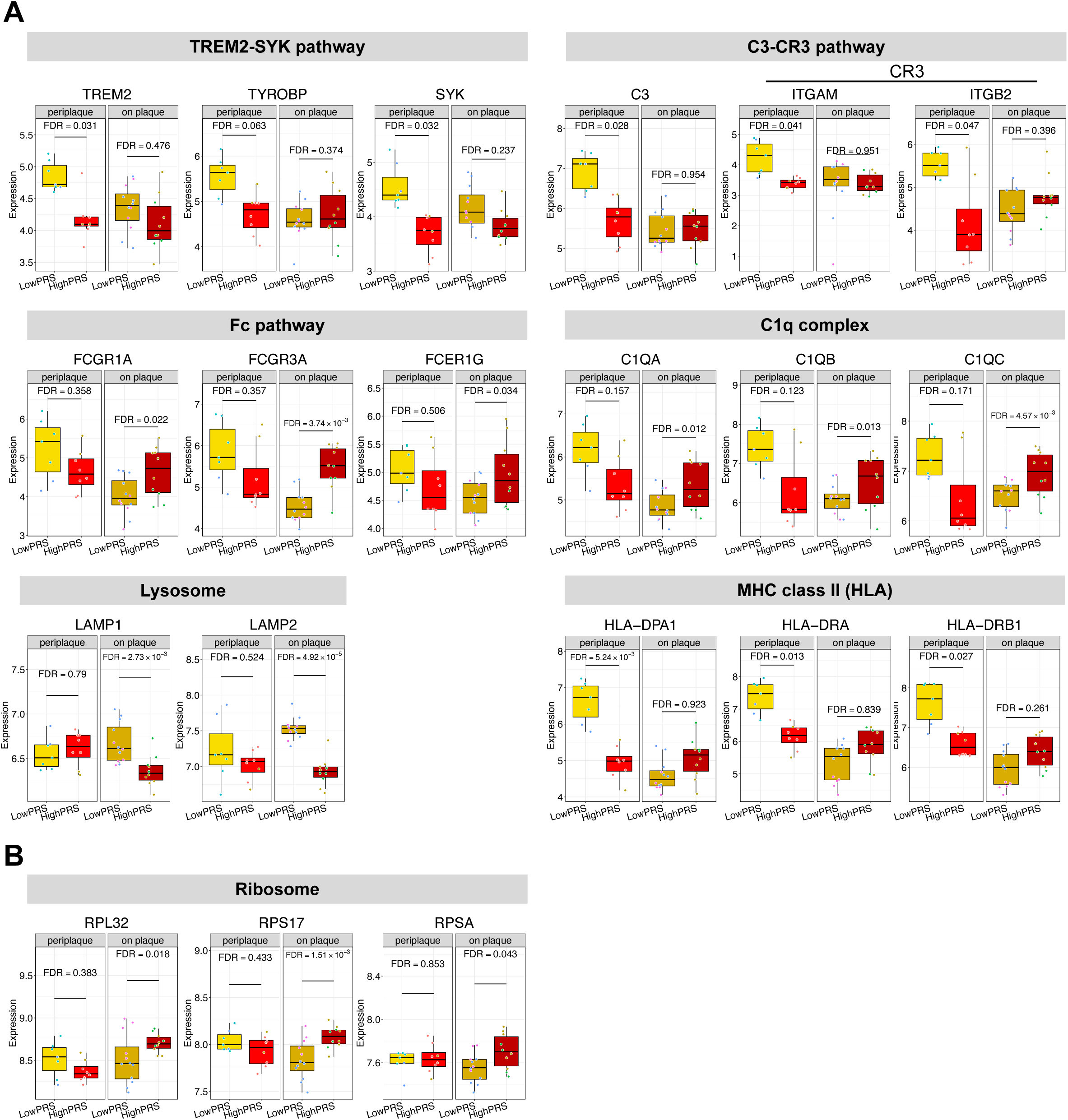
Expression levels of selected genes determined by spatial transcriptomics. (A) Box plots showing the expression of representative genes in Figure 6A. (B) Box plots of ribosomal gene expression.

**Figure S6.**
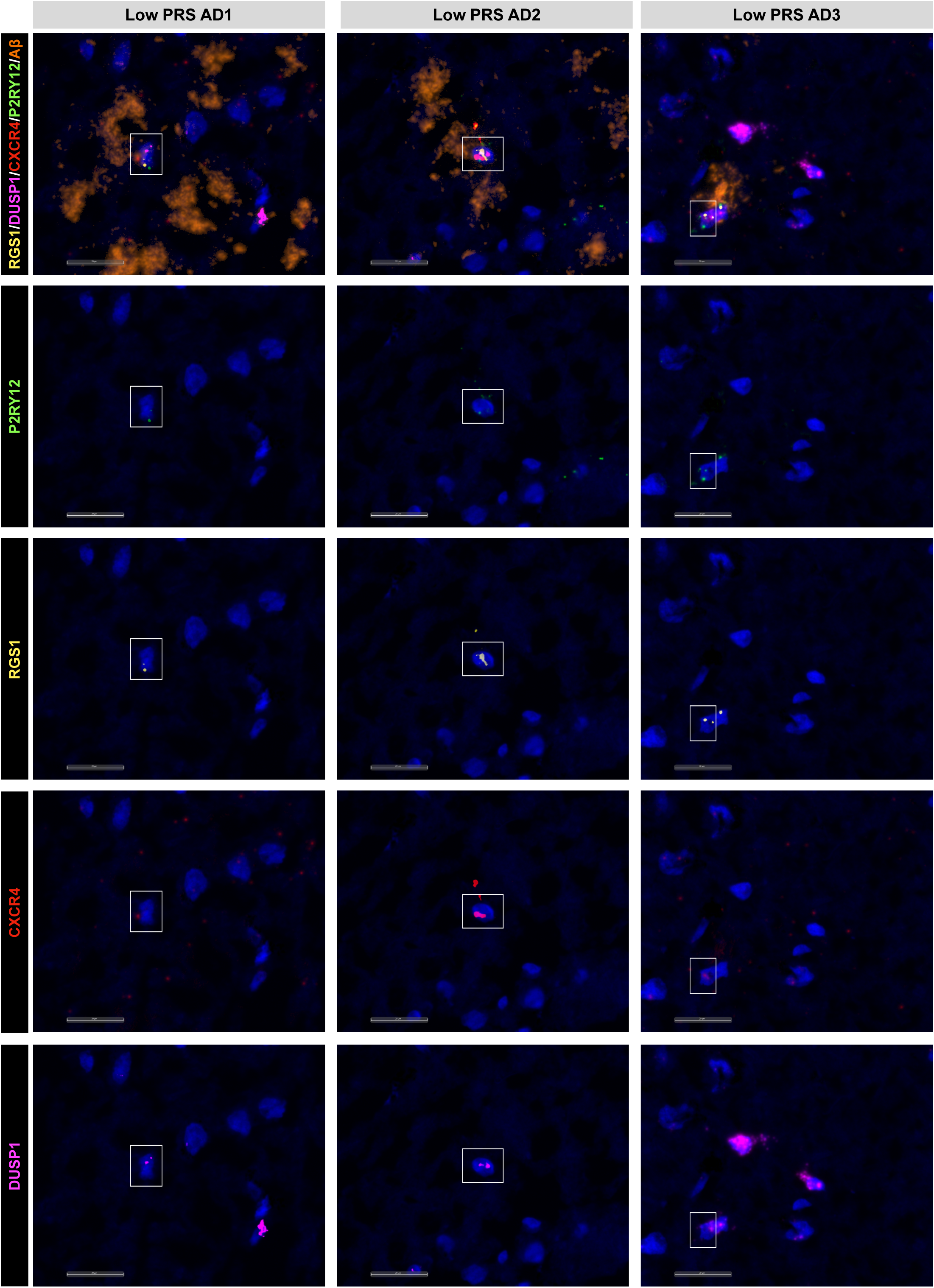
Immunohistochemistry for Aβ and RNAscope in situ hybridization for marker genes. Representative images correspond to postmortem frontal cortex tissue sections from low-PRS AD patients shown in Figure 7F. Scale bars: 20 μm.

